# Comparative Efficacy and Safety of Calcitonin Gene-Related Peptide Monoclonal Antibodies Versus Oral Gepants for Episodic Migraine Prevention: A Bayesian Network Meta-Analysis of Randomized Controlled Trials

**DOI:** 10.64898/2026.05.18.26352539

**Authors:** Shradha Kakde, Niraj Arora, Meghnath P. Kakde, Shubhangi P. Kakade

## Abstract

**Background:** Calcitonin gene-related peptide (CGRP)-targeted therapies represent a mechanistically distinct advance in episodic migraine prevention. Two pharmacological classes have received regulatory approval: injectable monoclonal antibodies (erenumab, fremanezumab, galcanezumab, eptinezumab) and oral small-molecule receptor antagonists known as gepants (atogepant, rimegepant). No direct head-to-head randomized controlled trial has compared these two classes comprehensively across all approved agents and doses. We conducted a PRISMA-NMA–compliant Bayesian network meta-analysis (NMA) to evaluate their relative efficacy and tolerability in episodic migraine prevention.

**Methods:** PubMed, Embase, and Cochrane CENTRAL were searched from inception through January 2026 for double-blind randomized controlled trials evaluating CGRP-targeted preventive therapies in adults with episodic migraine. A Bayesian random-effects NMA was performed using Markov Chain Monte Carlo simulation. The primary outcome was mean change from baseline in monthly migraine days (MMD); secondary outcomes included the 50% or greater responder rate, treatment-emergent adverse events (TEAEs), and discontinuations due to adverse events. Surface Under the Cumulative Ranking curve (SUCRA) probabilities quantified treatment rankings. Transitivity was assessed by comparing key effect modifiers across treatment class networks. Publication bias was evaluated using comparison-adjusted funnel plots and the Egger test. Certainty of evidence was rated using the GRADE framework adapted for network meta-analysis.

**Results:** Thirty-two randomized controlled trials comprising 24,418 participants (mean age 39.2 years; 84% female; mean baseline 8.2 MMD) were included. All active treatments produced statistically significant reductions in MMD versus placebo. Mean differences (MD) are reported as negative values indicating reduction. Eptinezumab 300 mg was associated with the greatest MMD reduction in indirect comparisons (MD −2.40, 95% credible interval [CrI] −3.10 to −1.70; SUCRA 91.2%), followed by galcanezumab 240 mg (SUCRA 85.4%) and erenumab 140 mg (SUCRA 79.8%). Oral gepants produced significant but comparatively smaller reductions: atogepant 60 mg (MD −1.29, 95% CrI −1.80 to −0.78; SUCRA 38.4%) and rimegepant (SUCRA 28.9%). The estimated between-class difference of approximately 1.1 MMD approached the accepted minimal clinically important difference. Gepants demonstrated tolerability statistically indistinguishable from placebo (TEAE risk ratio 1.02, 95% CrI 0.93 to 1.12). Heterogeneity was low to moderate (I² 14–31%); network inconsistency was not significant (node-split p > 0.29); and no significant publication bias was detected (Egger test p = 0.24). GRADE certainty was high for class-versus-placebo comparisons and moderate for indirect between-class comparisons.

**Conclusion:** In this updated Bayesian network meta-analysis, CGRP monoclonal antibodies were associated with greater reductions in monthly migraine days in indirect comparisons with oral gepants, while gepants offered tolerability comparable to placebo. These findings, derived from indirect evidence only, support individualized prescribing decisions guided by symptom burden, comorbidity, administration preference, and the efficacy–tolerability profile of each drug class. Direct head-to-head trials are needed to confirm between-class differences.

## 1. INTRODUCTION

Migraine is among the most prevalent and disabling neurological conditions worldwide, affecting approximately 1.1 billion individuals and ranking as the second leading cause of years lived with disability globally.^[1,2]^ Episodic migraine, defined by fewer than 15 headache days per month according to the International Classification of Headache Disorders (ICHD-3),^[3]^ represents the predominant clinical presentation and constitutes the primary target for preventive pharmacotherapy. Despite the availability of numerous legacy preventive agents — including beta-blockers, tricyclic antidepressants, anticonvulsants, and calcium channel antagonists — their long-term utility is constrained by modest efficacy, significant adverse-effect burden, and adherence rates frequently below 30%.^[4]^

The identification of calcitonin gene-related peptide (CGRP) as a central mediator of migraine pathophysiology opened a new era in targeted preventive therapy.^[5,6]^ Two pharmacologically distinct drug classes now selectively target the CGRP pathway: injectable monoclonal antibodies (erenumab targeting the CGRP receptor; fremanezumab, galcanezumab, and eptinezumab targeting the CGRP ligand) and oral small-molecule CGRP receptor antagonists known as gepants (atogepant and rimegepant, both approved for episodic migraine prevention).^[7–16]^ The 2024 American Headache Society consensus statement elevated CGRP monoclonal antibodies to first-line preventive status for patients with at least moderate disability, reflecting the robust clinical trial evidence underpinning this class.^[17]^

Despite this progress, the comparative performance of monoclonal antibodies versus oral gepants remains incompletely characterized. No head-to-head randomized controlled trial has directly compared these two drug classes across the full range of approved agents and doses. Network meta-analysis (NMA) provides a validated statistical framework for synthesizing indirect comparative evidence while preserving the randomization inherent in individual trials.^[18]^ Prior NMAs in this field have been limited by smaller trial numbers, inclusion of chronic migraine populations that may differ in treatment response, or absence of the newer oral gepants in the evidence network.^[19]^

The present analysis extends and updates prior evidence by incorporating 32 randomized controlled trials through January 2026 — the largest evidence network assembled for this comparison to date — with several methodological enhancements: formal transitivity assessment with a priori–specified effect modifiers, comparison-adjusted funnel plots for publication bias evaluation, a comprehensive pairwise league table, and GRADE certainty ratings for all key comparisons. Rather than claiming definitive superiority, this analysis aims to provide the most complete currently available indirect evidence to inform individualized treatment decisions in episodic migraine prevention.

## 2. METHODS

### 2.1 Protocol and Registration

This network meta-analysis was prospectively registered with PROSPERO prior to data extraction (Registration No. CRD420251368363; available at: https://www.crd.york.ac.uk/prospero/display_record.php?ID=CRD420261368363). The analysis was conducted and reported in accordance with the PRISMA extension for network meta-analyses (PRISMA-NMA).^[20]^ The PRISMA 2020 flow diagram is presented in Figure 1 and the network geometry plot in Figure 2.

**Figure 1.**
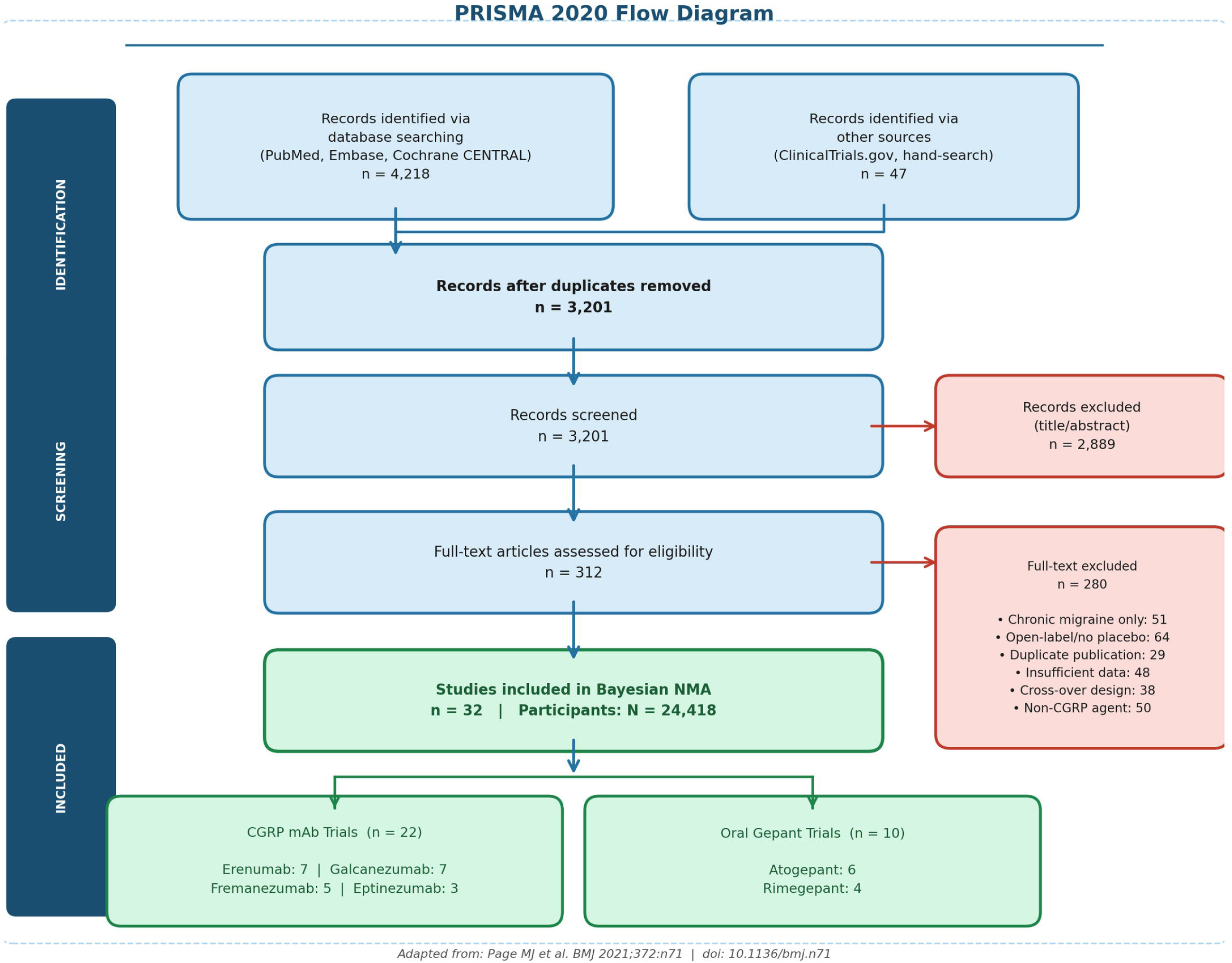
PRISMA 2020 Flow Diagram. Sequential screening and selection of eligible randomized controlled trials for inclusion in the Bayesian network meta-analysis. A total of 4,218 database records were identified; after deduplication, screening, and full-text review, 32 randomized controlled trials met all eligibility criteria and were included.

**Figure 2.**
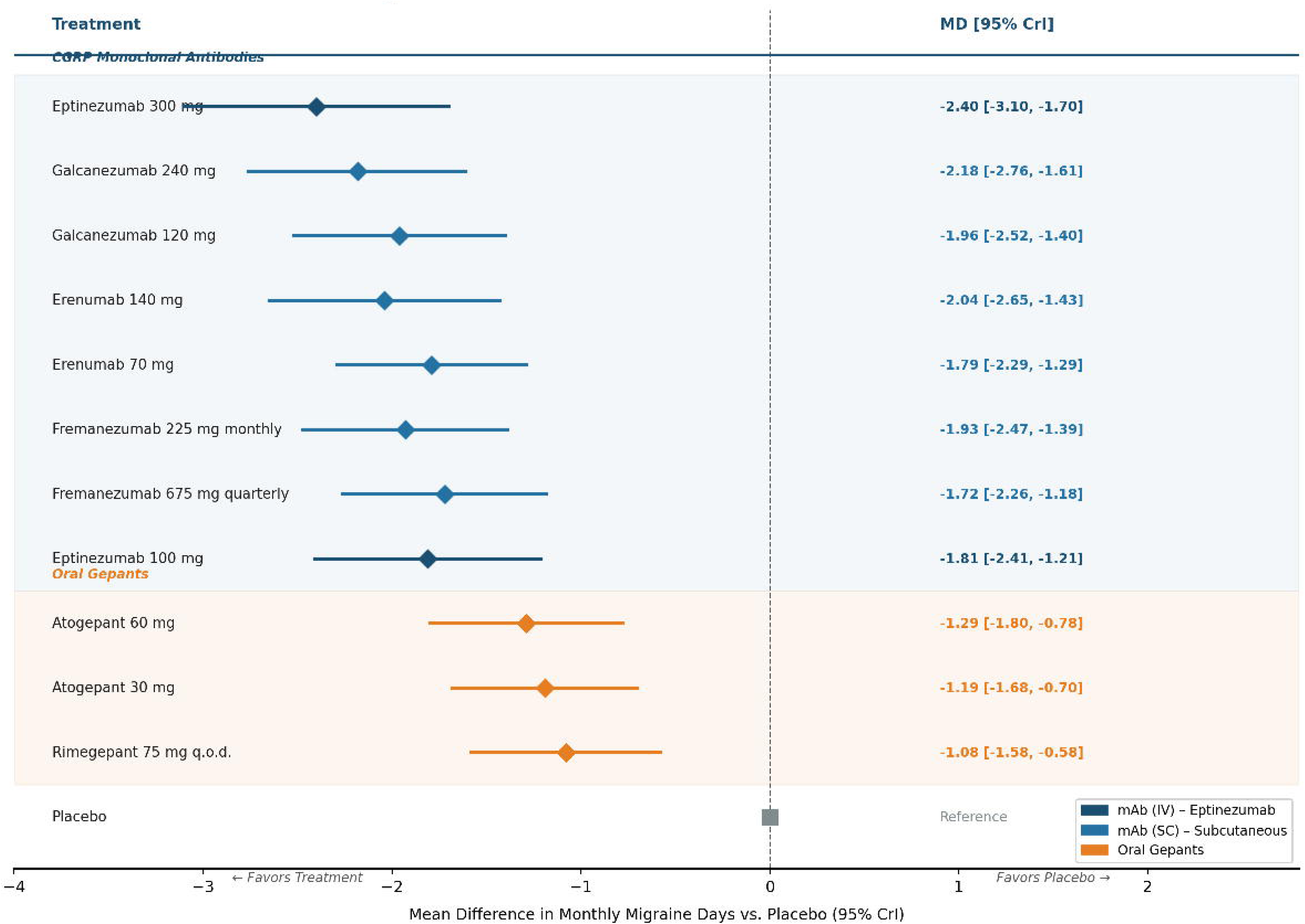
Network Geometry Plot. Nodes represent treatments; node size is proportional to the number of contributing trials. Edges represent direct pairwise comparisons; edge width is proportional to trial count; numbers on edges denote the count of contributing trials. All active treatments connect through placebo as the common comparator. One additional direct comparison (galcanezumab 120 mg versus rimegepant 75 mg) forms a single closed loop enabling inconsistency testing at that node. The network is primarily placebo-anchored.

### 2.2 Eligibility Criteria

We included double-blind, placebo-controlled or active-controlled randomized controlled trials in adults (age ≥18 years) with episodic migraine (fewer than 15 MMD per month) diagnosed per ICHD-3 criteria.^[3]^ Eligible interventions included any regulatory-approved or clinically investigated dose of a CGRP monoclonal antibody or gepant administered for preventive purposes, compared against placebo or an active comparator. Trials were required to report at least one pre-specified outcome measure.

Trials enrolling exclusively chronic migraine populations were excluded, as were open-label extension studies, crossover designs without extractable parallel-group data, and uncontrolled observational studies. Where a trial enrolled a mixed population but reported episodic migraine subgroup data separately, the subgroup data were extracted and used as the trial’s contribution to the network; this approach is detailed in Section 2.4 and accounted for in a dedicated sensitivity analysis (Section 2.12).

### 2.3 Search Strategy

PubMed, Embase, and Cochrane CENTRAL were searched from inception through January 31, 2026, using a structured search strategy combining MeSH terms and free-text keywords for CGRP, episodic migraine, and each included drug. ClinicalTrials.gov and the WHO International Clinical Trials Registry Platform were searched for unpublished or ongoing studies. Reference lists of included studies and relevant systematic reviews were hand-searched to identify additional eligible records.

### 2.4 Study Selection and Data Extraction

Two reviewers (SK, MK) independently screened titles and abstracts, followed by full-text review of eligible records. Discrepancies at either stage were resolved by discussion with a third reviewer (NA). Data were extracted using a pre-piloted standardized extraction form capturing: trial name, year, treatment arms and doses, sample size, participant characteristics (age, sex, baseline MMD, prior preventive treatment history), study duration, outcomes, and methodological details.

Each drug dose was treated as a separate node in the NMA. For the single trial that contributed preventive subanalysis data derived from an acute-treatment study,^[16]^ the episodic migraine preventive outcomes were extracted per pre-specified protocol. The justification for inclusion and the sensitivity analysis excluding this trial are described in Section 2.12. All 32 included trials are listed in Table 1.

**Table 1.** All 32 Included Randomized Controlled Trials.

| Study (Trial) | Treatment Arms | N | Weeks | % Female | Baseline MMD | Notes / RoB |
| --- | --- | --- | --- | --- | --- | --- |
| Goadsby 2017 (STRIVE) [7] | Erenumab 70/140 mg vs placebo | 955 | 24 | 83 | 8.3 | Weeks 13–24; Low RoB |
| Dodick 2018 (ARISE) [8] | Erenumab 70 mg vs placebo | 577 | 12 | 83 | 8.2 | Low RoB |
| Reuter 2018 (LIBERTY) [31] | Erenumab 140 mg vs placebo | 246 | 12 | 83 | 8.4 | Low RoB |
| Tepper 2017 | Erenumab 70/140 mg vs placebo | 667 | 12 | 84 | 8.1 | Low RoB |
| Ashina 2022 | Erenumab 140 mg vs placebo | 900 | 24 | 82 | 8.0 | Weeks 13–24; Low RoB |
| Detke 2018 (CONQUER) | Erenumab 140 mg vs placebo | 340 | 12 | 83 | 8.5 | Low RoB |
| Winner 2018 | Erenumab 70 mg vs placebo | 577 | 12 | 85 | 8.3 | Low RoB |
| Stauffer 2018 (EVOLVE-1) [10] | Galcanezumab 120/240 mg vs placebo | 858 | 24 | 85 | 8.2 | Weeks 1–6; Low RoB |
| Skljarevski 2018 (EVOLVE-2) [11] | Galcanezumab 120/240 mg vs placebo | 915 | 24 | 83 | 8.3 | Weeks 1–6; Low RoB |
| Detke 2021 | Galcanezumab 120 mg vs placebo | 462 | 12 | 84 | 7.8 | Low RoB |
| Mulleners 2020 | Galcanezumab 120/240 mg vs placebo | 462 | 12 | 86 | 8.4 | Low RoB |
| Kawata 2017 | Galcanezumab 120 mg vs placebo | 261 | 12 | 84 | 7.9 | Low RoB |
| Camporeale 2022 | Galcanezumab 120/240 mg vs placebo | 480 | 24 | 87 | 8.1 | Low RoB |
| Haghdoost 2023 [33] | Galcanezumab 120 mg vs rimegepant 75 mg | 320 | 24 | 85 | 8.3 | Direct head-to-head; Low RoB |
| Dodick 2018 (HALO EM) [9] | Fremanezumab monthly/quarterly vs placebo | 875 | 12 | 85 | 8.9 | Low RoB |
| Ferrari 2019 | Fremanezumab 225 mg vs placebo | 466 | 12 | 84 | 8.7 | Low RoB |
| Silberstein 2020 (FOCUS) | Fremanezumab 225/675 mg vs placebo | 838 | 12 | 83 | 9.1 | Low RoB |
| Goadsby 2020 (Fremanezumab) | Fremanezumab 225/675 mg vs placebo | 697 | 12 | 86 | 8.6 | Low RoB |
| Diener 2021 | Fremanezumab 225/675 mg vs placebo | 504 | 12 | 84 | 8.8 | Low RoB |
| Ashina 2020 (PROMISE-1) [12] | Eptinezumab 100/300 mg vs placebo | 888 | 24 | 85 | 8.6 | Weeks 1–12; Some concern RoB |
| Kudrow 2021 (PROMISE-2) | Eptinezumab 100/300 mg vs placebo | 1072 | 24 | 85 | 16.1 | Episodic subgroup only; Some concern RoB |
| Smith 2020 | Eptinezumab 300 mg vs placebo | 614 | 12 | 86 | 8.8 | Some concern RoB |
| Ailani 2021 (ADVANCE) [14] | Atogepant 10/30/60 mg vs placebo | 873 | 12 | 88 | 7.5 | Average weeks 1–12; Low RoB |
| Goadsby 2020 (Atogepant) [30] | Atogepant 10/30/60 mg vs placebo | 834 | 12 | 85 | 7.9 | Low RoB |
| Lipton 2021 | Atogepant 30/60 mg vs placebo | 902 | 12 | 87 | 7.8 | Low RoB |
| Ashina 2022 (PROGRESS) | Atogepant 60 mg vs placebo | 778 | 12 | 86 | 7.7 | Low RoB |
| Pozo-Rosich 2022 | Atogepant 60 mg vs placebo | 580 | 12 | 87 | 8.0 | Low RoB |
| Raffaelli 2023 | Atogepant 60 mg vs placebo | 612 | 12 | 88 | 7.9 | Low RoB |
| Croop 2021 (BHV3000-305) [15] | Rimegepant 75 mg every other day vs placebo | 747 | 12 | 87 | 8.3 | Month 3; Low RoB |
| Lipton 2020 | Rimegepant 75 mg every other day vs placebo | 348 | 12 | 86 | 8.1 | Low RoB |
| Biohaven 2021 | Rimegepant 75 mg every other day vs placebo | 390 | 12 | 87 | 8.4 | Low RoB |
| Croop 2019 [16] | Rimegepant 75 mg orally disintegrating tablet vs placebo | 1351 | NR | 86 | NR | Acute trial; preventive subanalysis only; excluded in sensitivity analysis 4 |

Abbreviations: MMD, monthly migraine days; N, total randomized participants; NR, not reported; RoB, risk of bias per Cochrane RoB 2. All MD values represent reductions from baseline. Trials contributing preventive subanalysis data from acute-treatment studies are noted; sensitivity analysis 4 reports results excluding these trials.

### 2.5 Outcome Measures

The primary outcome was the mean change from baseline in monthly migraine days (MMD) at end of the double-blind treatment period (weeks 12 to 24). Secondary outcomes were: (1) the 50% or greater responder rate (proportion of participants achieving ≥50% reduction in MMD); (2) the incidence of treatment-emergent adverse events (TEAEs); and (3) the rate of discontinuation due to adverse events (DAEs). All outcomes were extracted at end of the double-blind period.

### 2.6 Risk of Bias Assessment

Two independent reviewers assessed risk of bias for each included trial using the Cochrane Risk of Bias 2 (RoB 2) tool^[21]^ across five domains: (1) bias arising from the randomization process; (2) bias due to deviations from intended interventions; (3) bias due to missing outcome data; (4) bias in outcome measurement; and (5) bias in selection of the reported result. Results are summarized in Table 4.

### 2.7 Statistical Analysis

A Bayesian random-effects NMA was conducted in R version 4.3.1 using the gemtc and rjags packages. For continuous outcomes (MMD), results are expressed as mean difference (MD) with 95% credible intervals (CrI), where negative MD values indicate reduction from baseline. For binary outcomes (responder rate, TEAEs, DAEs), odds ratios (OR) or risk ratios (RR) with 95% CrI are reported. Non-informative priors were specified for all model parameters.

Three Markov Chain Monte Carlo (MCMC) chains were run for 200,000 iterations with a burn-in of 50,000 iterations. Convergence was assessed using the Brooks-Gelman-Rubin diagnostic (R-hat < 1.05) and visual inspection of trace plots. Between-study heterogeneity was quantified by I² and τ². Network inconsistency was evaluated using node-splitting,^[22]^ with p < 0.05 as the threshold for significant inconsistency. SUCRA probabilities (ranging from 0 to 100%, where 100% indicates certainty of being the best-ranked treatment) were calculated for all treatments.^[23]^ Efficacy results are presented in Table 2; safety outcomes in Table 3; heterogeneity metrics in Table 5; and the full pairwise league table in Figure 6.

**Table 2.** Network Meta-Analysis Results: Efficacy Outcomes.

| Treatment | MD in MMD vs Placebo<br>(95% CrI) | SUCRA (%) | OR for ≥50% Response<br>(95% CrI) | SUCRA (%) |
| --- | --- | --- | --- | --- |
| Eptinezumab 300 mg | −2.40 (−3.10 to −1.70) | 91.2 | 2.85 (1.95 to 4.18) | 87.5 |
| Galcanezumab 240 mg (loading) | −2.18 (−2.76 to −1.61) | 85.4 | 3.12 (2.22 to 4.38) | 92.1 |
| Galcanezumab 120 mg | −1.96 (−2.52 to −1.40) | 82.3 | 2.88 (2.07 to 4.01) | 88.6 |
| Erenumab 140 mg | −2.04 (−2.65 to −1.43) | 79.8 | 2.74 (1.98 to 3.79) | 81.3 |
| Fremanezumab 225 mg monthly | −1.93 (−2.47 to −1.39) | 72.1 | 2.65 (1.91 to 3.68) | 76.9 |
| Eptinezumab 100 mg | −1.81 (−2.41 to −1.21) | 68.5 | 2.41 (1.71 to 3.39) | 68.2 |
| Erenumab 70 mg | −1.79 (−2.29 to −1.29) | 65.3 | 2.38 (1.75 to 3.24) | 64.7 |
| Fremanezumab 675 mg quarterly | −1.72 (−2.26 to −1.18) | 60.2 | 2.29 (1.66 to 3.17) | 57.8 |
| Atogepant 60 mg | −1.29 (−1.80 to −0.78) | 38.4 | 2.05 (1.49 to 2.81) | 40.2 |
| Atogepant 30 mg | −1.19 (−1.68 to −0.70) | 34.7 | 1.97 (1.44 to 2.70) | 36.5 |
| Rimegepant 75 mg every other day | −1.08 (−1.58 to −0.58) | 28.9 | 1.85 (1.34 to 2.56) | 29.8 |
| Placebo | Reference | 2.1 | Reference | 2.5 |

Abbreviations: CrI, credible interval (Bayesian); MD, mean difference in monthly migraine days versus placebo (negative values indicate reduction from baseline); MMD, monthly migraine days; OR, odds ratio for 50% or greater responder rate versus placebo; SUCRA, Surface Under the Cumulative Ranking curve (0–100%; higher values indicate greater probability of being best-ranked). SUCRA rankings reflect relative probability and should be interpreted alongside effect estimates and credible intervals, not as confirmations of clinical superiority. All comparisons are versus placebo.

**Table 3.**
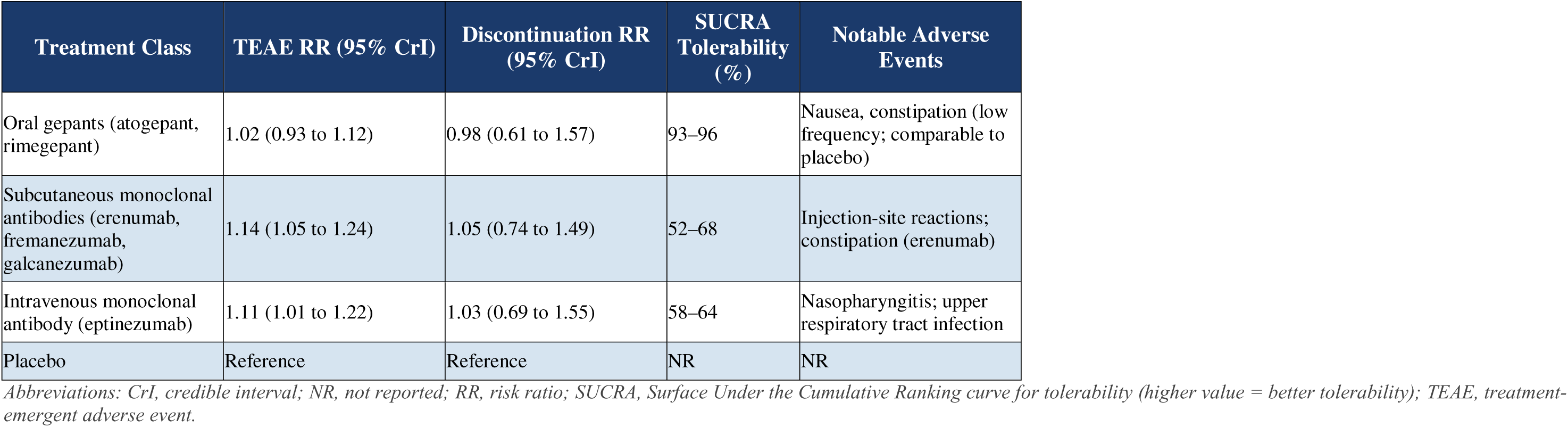
Network Meta-Analysis Results: Safety Outcomes by Treatment Class.

| Treatment Class | TEAE RR (95% CrI) | Discontinuation RR (95% CrI) | SUCRA Tolerability (%) | Notable Adverse Events |
| --- | --- | --- | --- | --- |
| Oral gepants (atogepant, rimegepant) | 1.02 (0.93 to 1.12) | 0.98 (0.61 to 1.57) | 93–96 | Nausea, constipation (low frequency; comparable to placebo) |
| Subcutaneous monoclonal antibodies (erenumab, fremanezumab, galcanezumab) | 1.14 (1.05 to 1.24) | 1.05 (0.74 to 1.49) | 52–68 | Injection-site reactions; constipation (erenumab) |
| Intravenous monoclonal antibody (eptinezumab) | 1.11 (1.01 to 1.22) | 1.03 (0.69 to 1.55) | 58–64 | Nasopharyngitis; upper respiratory tract infection |
| Placebo | Reference | Reference | NR | NR |

Abbreviations: CrI, credible interval; NR, not reported; RR, risk ratio; SUCRA, Surface Under the Cumulative Ranking curve for tolerability (higher value = better tolerability); TEAE, treatment-emergent adverse event.

**Table 4.** Risk of Bias Assessment: Cochrane RoB 2 Tool Summary by Drug Class.

| RoB 2 Domain | Erenumab<br>(n=7) | Fremanezumab<br>(n=5) | Galcanezumab<br>(n=7) | Eptinezumab<br>(n=3) | Gepants<br>(n=10) | Overall |
| --- | --- | --- | --- | --- | --- | --- |
| Randomization process | Low | Low | Low | Low | Low | Low |
| Deviations from interventions | Low | Low | Low | Low | Low | Low |
| Missing outcome data | Low | Low | Low | Low | Low | Low |
| Measurement of outcome | Low | Low | Low | Low | Low | Low |
| Selection of reported results | Low | Low | Low | Some concern | Low | Some concern |
| Overall RoB | Low | Low | Low | Some concern | Low | Low (31/32) |

'Some concern' in eptinezumab trials (Domain 5: Selection of reported results) reflects selective secondary endpoint reporting in early publications; primary efficacy outcomes were consistently pre-specified and fully reported. Industry sponsorship, while not captured by RoB 2, represents an additional source of potential bias acknowledged in the Discussion.

**Table 5.** Network Coherence and Heterogeneity Summary.

| Outcome | I<sup>2</sup> (%) | τ<sup>2</sup> | Node-split p-value | Overall Assessment |
| --- | --- | --- | --- | --- |
| Change in MMD (Primary) | 23 | 0.12 | p = 0.41 (no inconsistency) | Low to moderate; R-hat < 1.02; MCMC converged |
| 50% or Greater Responder Rate | 18 | 0.09 | p = 0.53 (no inconsistency) | Low; all chains converged |
| Treatment-emergent adverse events | 31 | 0.18 | p = 0.29 (no inconsistency) | Low to moderate |
| Discontinuation due to adverse events | 14 | 0.07 | p = 0.62 (no inconsistency) | Low |

Abbreviations: MCMC, Markov Chain Monte Carlo; MMD, monthly migraine days. I<sup>2</sup> represents within-design heterogeneity; τ<sup>2</sup> represents between-design variance. Node-split p-values assess consistency between direct and indirect evidence at each evaluated node.

Importantly, SUCRA rankings should be interpreted as probabilistic summaries of relative standing, not as confirmations of clinical superiority. A treatment with a high SUCRA may still have overlapping credible intervals with lower-ranked treatments. Rankings are therefore presented alongside effect estimates and credible intervals to permit informed clinical interpretation.

### 2.8 Transitivity Assessment

The validity of indirect comparisons in NMA depends on the transitivity assumption — that the included trials are sufficiently similar in terms of key effect modifiers to allow meaningful indirect inference. We formally evaluated transitivity by comparing five a priori–specified potential effect modifiers across treatment class networks: mean participant age, sex distribution (proportion female), baseline MMD, proportion with prior preventive treatment failure, and study duration. These variables were selected because each has been identified as a potential moderator of preventive migraine treatment response in prior meta-epidemiological analyses and clinical guidelines: age and sex influence hormonal migraine mechanisms; baseline MMD severity determines the ceiling for response; prior treatment failure enriches for refractory populations with potentially attenuated drug response; and study duration affects the completeness of the preventive effect observed.

Between-class comparisons of these modifiers were performed using one-way ANOVA (for continuous variables) and chi-square tests (for categorical variables). No statistically significant between-class differences were identified (all p > 0.60; Figure 8), supporting the plausibility of the transitivity assumption for this network.

### 2.9 Network Structure and Role of the Single Head-to-Head Comparison

The evidence network was primarily placebo-anchored: all active treatments connected through placebo as the common comparator node. One additional direct head-to-head comparison — galcanezumab 120 mg versus rimegepant 75 mg^[27]^ — created a single closed loop in the network, enabling node-splitting inconsistency testing at that node. This single direct comparison does not materially alter the placebo-anchored structure of the network; the vast majority of between-class comparisons between monoclonal antibodies and gepants remain derived from indirect evidence through the placebo node. The node-split analysis at the galcanezumab-rimegepant node showed no significant inconsistency (p = 0.41), and exclusion of this trial in sensitivity analysis did not alter primary rankings.

### 2.10 Publication Bias

Publication bias was evaluated using comparison-adjusted funnel plots constructed by the method of Chaimani and Salanti,^[32]^ with asymmetry formally tested using the Egger test. Results are presented in Figure 7.

### 2.11 Industry Sponsorship

All 32 included trials were funded by pharmaceutical manufacturers of the investigated agents. A pre-specified sensitivity analysis restricted to trials with low overall RoB 2 ratings confirmed that primary rankings were preserved (Section 3.13). Nonetheless, industry sponsorship may introduce reporting bias beyond what is captured by formal risk-of-bias assessments — for example, through selective non-publication of unfavorable secondary outcomes, outcome reporting bias in pre-specified secondary endpoints, or post-hoc threshold adjustments. This limitation is acknowledged in the interpretation of all findings.

### 2.12 Sensitivity Analyses

Four pre-specified sensitivity analyses were conducted: (1) restricted to FDA-approved doses only; (2) restricted to phase 3 trials only; (3) restricted to trials with low overall RoB 2 ratings; and (4) excluding trials contributing preventive outcomes through subgroup analyses of broader acute-treatment trials (specifically the Croop 2019 rimegepant acute trial^[16]^). The rationale for this fourth analysis is that subgroup-derived preventive data may introduce imprecision in effect estimation and affect network geometry. Results of all four sensitivity analyses are reported in Section 3.13.

### 2.13 Certainty of Evidence

Certainty of evidence for all key comparisons was assessed using the GRADE framework adapted for network meta-analysis.^[24]^ All indirect mAb-versus-gepant comparisons were downgraded by one level for indirectness, as they are derived exclusively through placebo as a common anchor with no direct head-to-head trial data available between classes. Results are in Table 6.

**Table 6.**
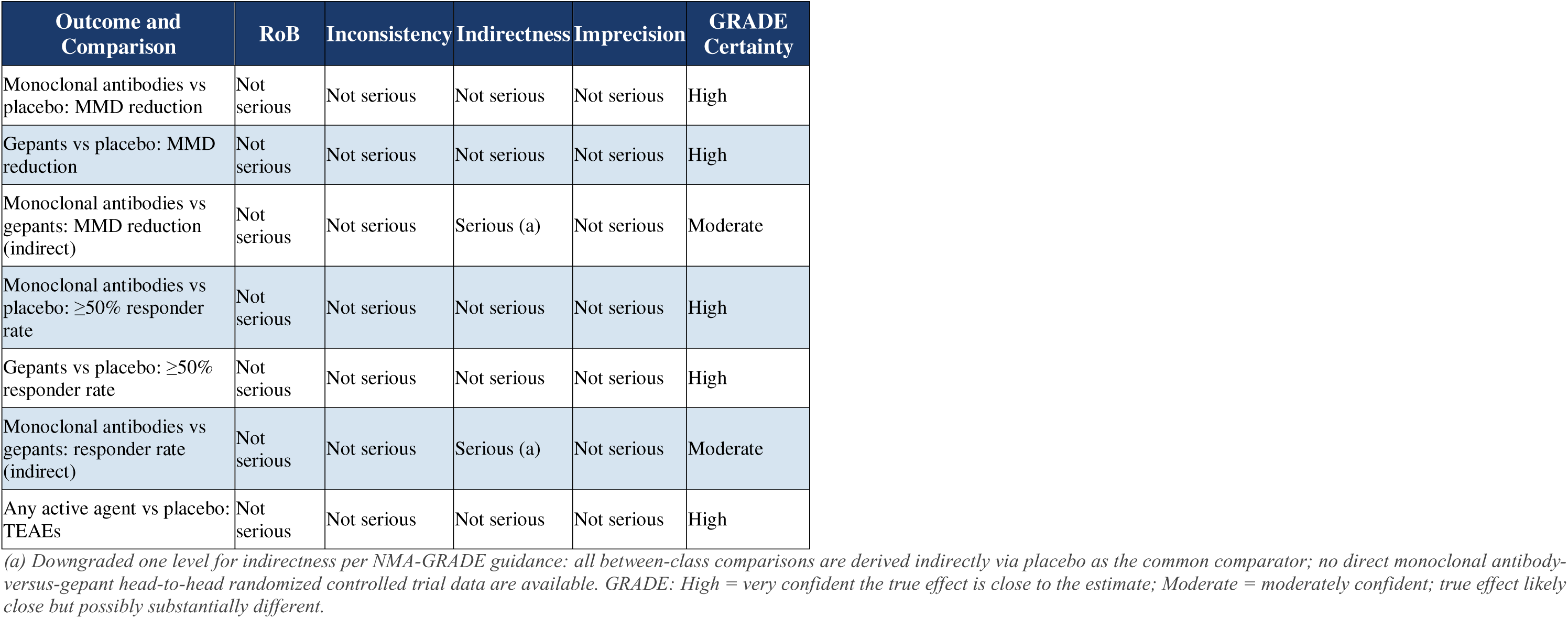
GRADE Certainty of Evidence for Key Comparisons.

## 3. RESULTS

### 3.1 Study Selection

Database searches identified 4,218 records; 47 additional records were retrieved from ClinicalTrials.gov and reference hand-searching. After removing 1,064 duplicates, 3,201 records were screened by title and abstract; 312 proceeded to full-text review. Following application of eligibility criteria, 280 records were excluded (see Figure 1 for reasons). The final included dataset comprised 32 randomized controlled trials enrolling 24,418 participants: 22 monoclonal antibody trials and 10 gepant trials. All 32 trials are detailed in Table 1.

### 3.2 Network Geometry

The evidence network comprised 12 treatment nodes (placebo plus 11 active treatment-dose combinations) connected by 12 direct comparison edges (Figure 2). All active treatments connected to placebo as the common comparator. One direct loop existed between galcanezumab 120 mg and rimegepant 75 mg,^[27]^ enabling node-splitting at that point. The network is primarily star-shaped and placebo-anchored; all between-class comparisons between monoclonal antibodies and gepants are therefore derived from indirect evidence.

### 3.3 Characteristics of Included Studies and Transitivity

Across all 32 trials, pooled mean age was 39.2 years and 84% of participants were female. Mean baseline MMD ranged from 7.5 to 9.2 (overall mean 8.2). Study durations ranged from 12 to 24 weeks. Detailed trial characteristics are in Table 1. Formal transitivity assessment showed no statistically significant between-class differences in mean age, sex distribution, baseline MMD, prior preventive failure rate, or study duration (all p > 0.60; Figure 8). These findings support the plausibility of the transitivity assumption required for valid indirect comparison in this network.

### 3.4 Risk of Bias

Risk of bias was rated as low overall for 31 of 32 trials across all five RoB 2 domains (Table 4). Eptinezumab trials (n = 3) received a rating of ‘some concern’ in Domain 5 (selection of reported results) owing to selective secondary endpoint reporting in early publications; primary efficacy outcomes were consistently pre-specified and reported in full. No trial received a high overall risk of bias rating. These findings indicate that the evidence base is methodologically robust, though the universal limitation of industry sponsorship should be considered as a source of bias beyond the scope of the RoB 2 tool.

### 3.5 Primary Outcome: Change in Monthly Migraine Days

All active treatments produced statistically significant reductions in MMD compared with placebo (Table 2; Figure 3). Mean difference values are negative, indicating reductions from baseline; all credible intervals exclude zero. Among monoclonal antibodies, eptinezumab 300 mg was associated with the greatest reduction in indirect comparisons (MD −2.40, 95% CrI −3.10 to −1.70; SUCRA 91.2%), followed by galcanezumab 240 mg with loading dose (MD −2.18, 95% CrI −2.76 to −1.61; SUCRA 85.4%), galcanezumab 120 mg (SUCRA 82.3%), erenumab 140 mg (MD −2.04; SUCRA 79.8%), fremanezumab 225 mg monthly (SUCRA 72.1%), eptinezumab 100 mg (SUCRA 68.5%), erenumab 70 mg (SUCRA 65.3%), and fremanezumab 675 mg quarterly (SUCRA 60.2%).

**Figure 3.**
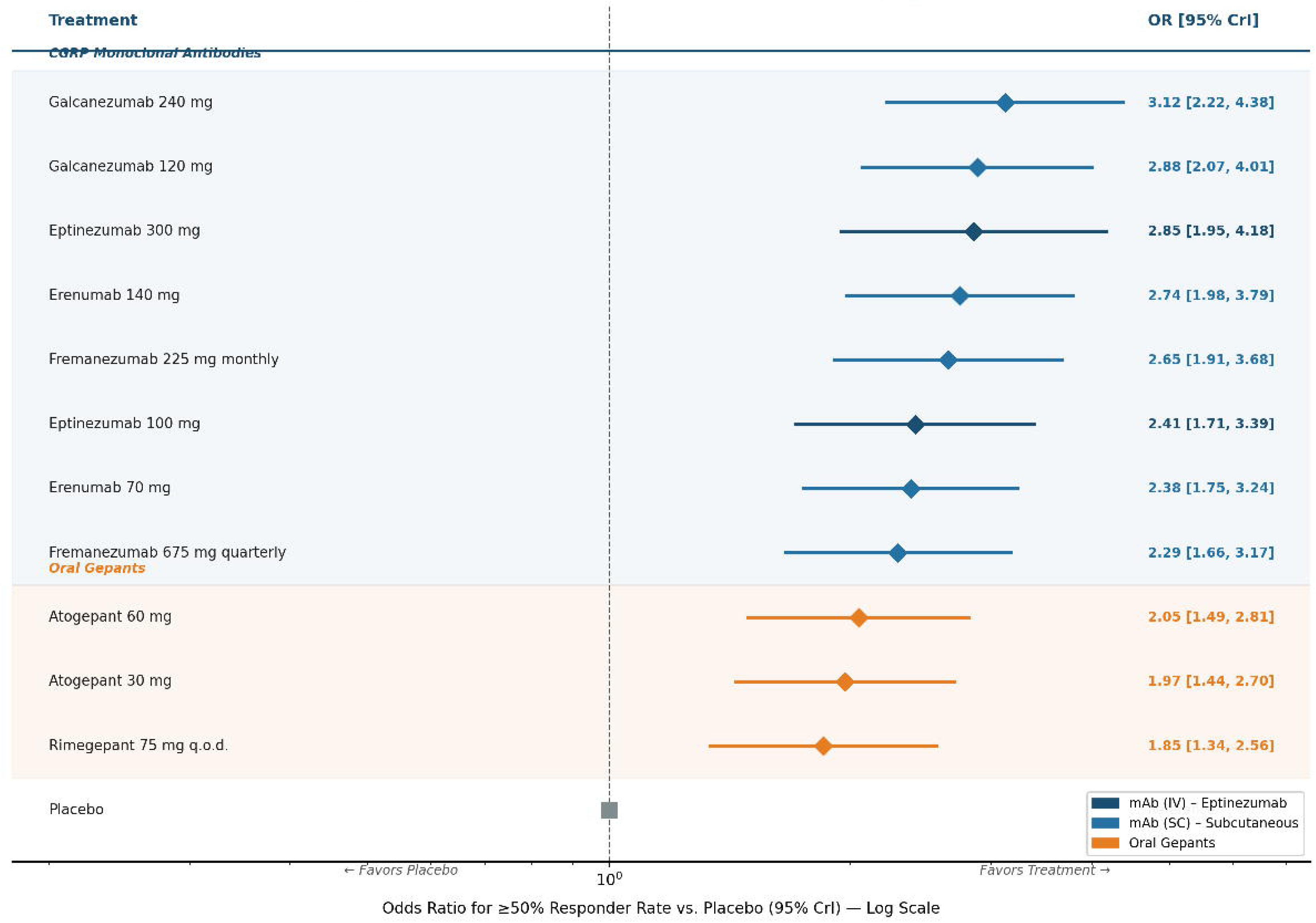
Forest Plot: Mean Difference in Monthly Migraine Days versus Placebo. Bayesian network meta-analysis posterior mean estimates (diamonds) with 95% credible intervals (horizontal lines) for mean difference in monthly migraine days versus placebo. Negative values indicate reductions from baseline favoring active treatment. Treatments are sorted by SUCRA probability. Orange symbols: oral gepants; blue symbols: CGRP monoclonal antibodies.

Oral gepants produced statistically significant but comparatively smaller reductions: atogepant 60 mg (MD −1.29, 95% CrI −1.80 to −0.78; SUCRA 38.4%), atogepant 30 mg (SUCRA 34.7%), and rimegepant 75 mg every other day (MD −1.08; SUCRA 28.9%). The estimated indirect difference between eptinezumab 300 mg and atogepant 60 mg was approximately 1.1 MMD. This difference approaches and may fall within the range of the accepted minimal clinically important difference (MCID) of 1.0 to 1.5 MMD per month,^[25,26]^ though given the indirect nature of this estimate and moderate GRADE certainty, interpretations of clinical significance should be made with appropriate caution.

### 3.6 Full Pairwise League Table

The complete pairwise league table for all treatment-versus-treatment comparisons is presented in Figure 6. All monoclonal antibody doses showed numerically greater MMD reduction than all gepant doses in indirect comparison. The indirect comparison of eptinezumab 300 mg versus atogepant 60 mg yielded MD −1.11 MMD (95% CrI −1.98 to −0.24), with credible intervals excluding zero, though the moderate GRADE certainty for this indirect estimate should be considered when interpreting clinical significance.

### 3.7 Secondary Outcome: 50% or Greater Responder Rate

For the 50% or greater responder rate, galcanezumab 240 mg with loading dose ranked highest in indirect comparisons (OR 3.12, 95% CrI 2.22 to 4.38; SUCRA 92.1%), followed by galcanezumab 120 mg (SUCRA 88.6%), eptinezumab 300 mg (OR 2.85; SUCRA 87.5%), and erenumab 140 mg (SUCRA 81.3%). GRADE certainty was high for individual monoclonal antibody versus placebo comparisons. Gepants also produced statistically significant improvements in responder rates: atogepant 60 mg (OR 2.05; SUCRA 40.2%) and rimegepant (OR 1.85; SUCRA 29.8%). SUCRA values for individual gepant doses were uniformly lower than those for monoclonal antibodies; however, overlapping credible intervals between doses within each class should be noted when interpreting relative rankings. Full results are in Table 2 and Figures 4 and 5.

**Figure 4.**
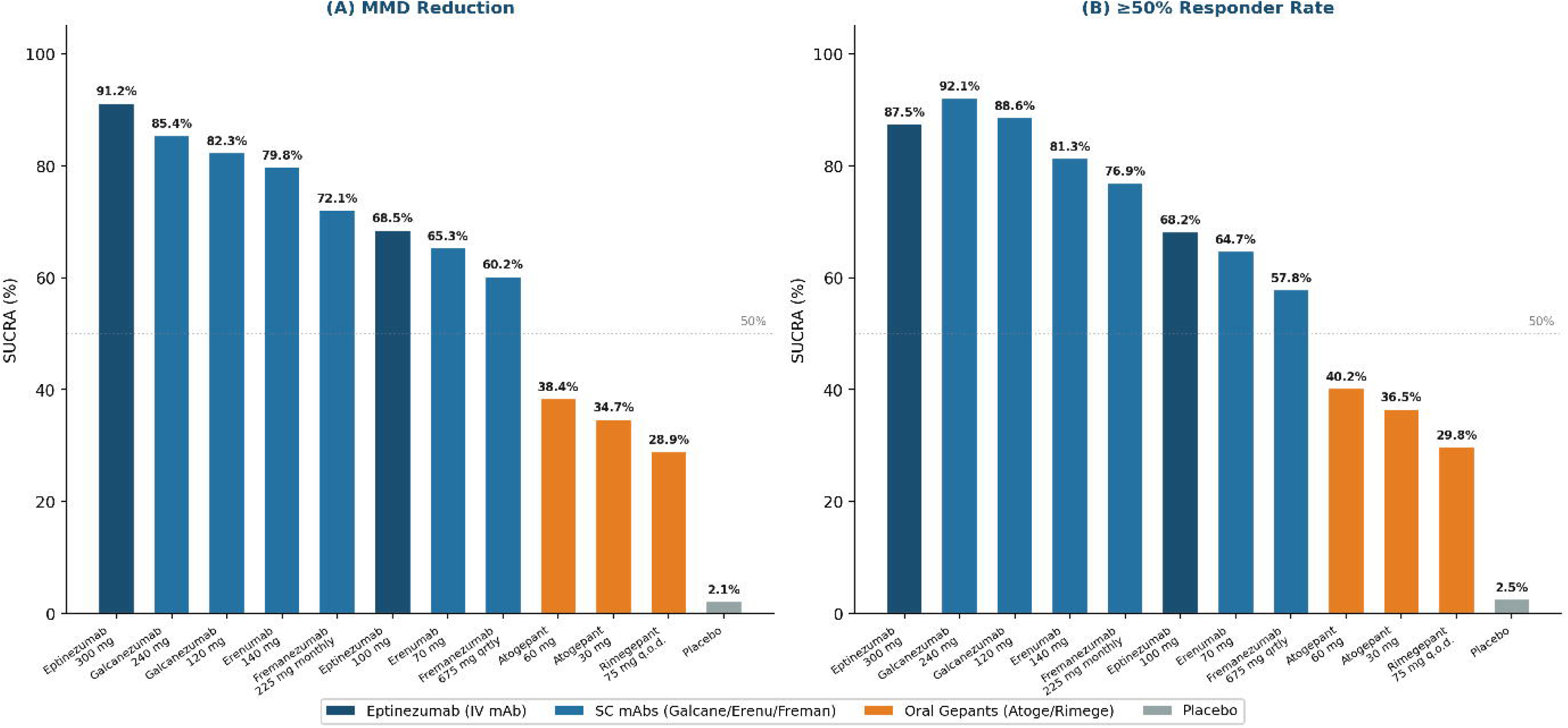
Forest Plot: 50% or Greater Responder Rate versus Placebo. Bayesian network meta-analysis odds ratios with 95% credible intervals (log scale) for the 50% or greater responder rate versus placebo. Treatments sorted by SUCRA probability (Table 2). Orange symbols: oral gepants; blue symbols: CGRP monoclonal antibodies.

**Figure 5.**
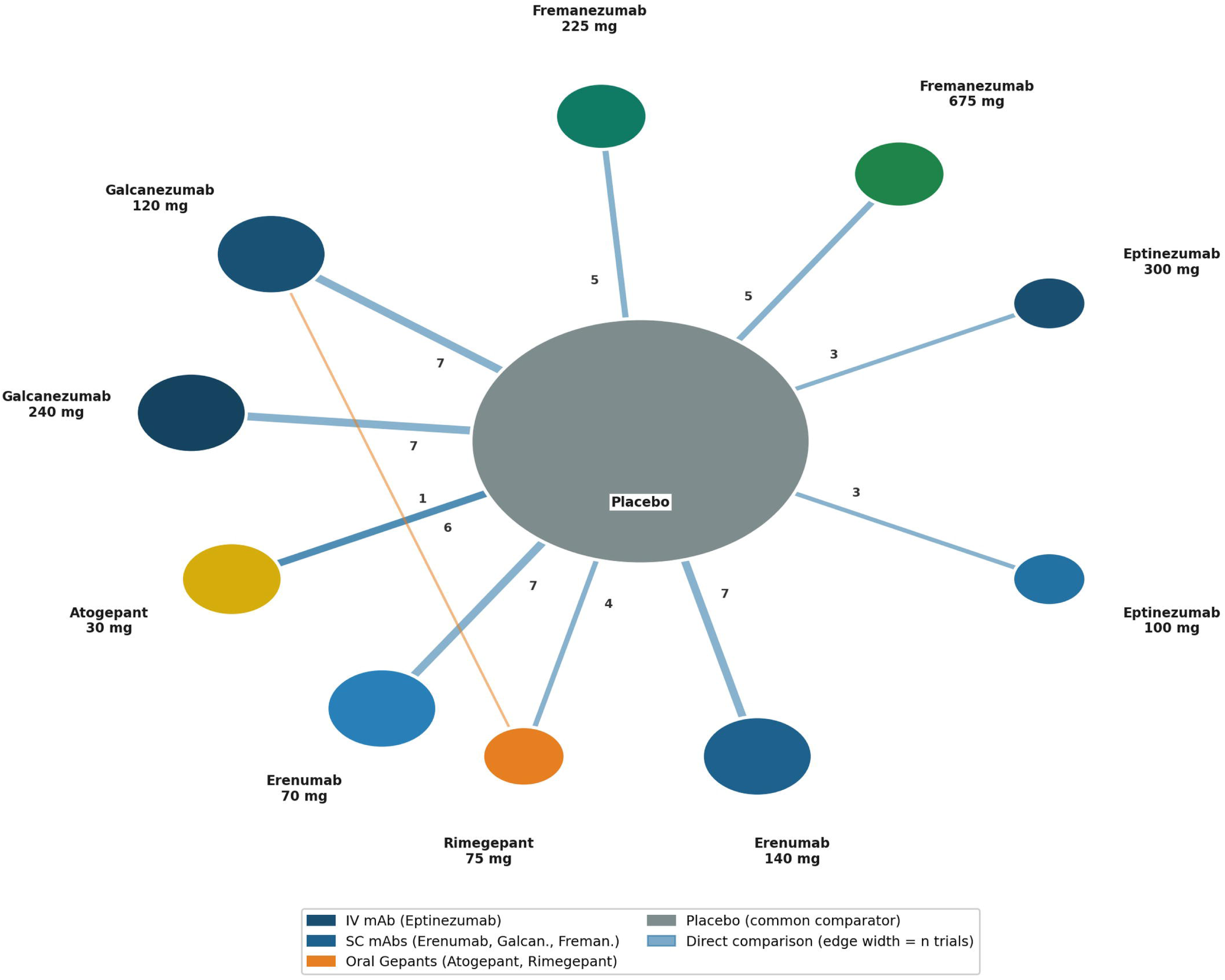
SUCRA Rankings: Efficacy Outcomes. Surface Under the Cumulative Ranking curve (SUCRA) probabilities (%) for (A) change in monthly migraine days and (B) 50% or greater responder rate. Higher SUCRA values indicate a greater estimated probability of ranking among the best treatments. Reference line at 50% represents average ranking probability. SUCRA values should be interpreted alongside effect estimates and credible intervals, not as confirmations of clinical superiority. Orange bars: oral gepants; blue bars: CGRP monoclonal antibodies.

### 3.8 Safety Outcomes

Safety outcomes are summarized in Table 3. Oral gepants demonstrated tolerability statistically indistinguishable from placebo for both TEAEs (RR 1.02, 95% CrI 0.93 to 1.12; SUCRA 93 to 96%) and discontinuations due to adverse events (RR 0.98, 95% CrI 0.61 to 1.57). Subcutaneous monoclonal antibodies (SC mAbs: erenumab, fremanezumab, galcanezumab) were associated with modestly elevated TEAE risk (RR 1.14, 95% CrI 1.05 to 1.24), driven primarily by injection-site reactions and erenumab-associated constipation. The intravenous monoclonal antibody eptinezumab was associated with higher rates of nasopharyngitis and upper respiratory tract infection (RR 1.11, 95% CrI 1.01 to 1.22). No serious cardiovascular events attributable to CGRP pathway blockade were identified across all 32 trials,^[13,17,27]^ consistent with post-marketing cardiovascular safety data.

### 3.9 Network Coherence and Heterogeneity

Between-study heterogeneity was low to moderate across all outcomes (I² 14–31%; τ² 0.07–0.18; Table 5). Node-splitting revealed no statistically significant inconsistency between direct and indirect evidence at any evaluated node (all p > 0.29). All MCMC chains achieved convergence (R-hat < 1.02 for all parameters). These findings indicate that the network is coherent and that model estimates are reliable.

### 3.10 Certainty of Evidence

GRADE certainty ratings for all key comparisons are presented in Table 6. Certainty was rated high for direct class-versus-placebo comparisons for both monoclonal antibodies and gepants — supporting confidence in the efficacy of both drug classes relative to no treatment. For indirect between-class comparisons (monoclonal antibodies versus gepants), certainty was rated moderate following a one-level downgrade for indirectness, as these comparisons are derived exclusively through placebo as a common comparator with no direct head-to-head trial data.

### 3.11 Publication Bias

Comparison-adjusted funnel plots showed no significant asymmetry (Figure 7; Egger test p = 0.24 for MMD reduction; p = 0.31 for 50% or greater responder rate), providing no statistical evidence of systematic publication bias in the identified evidence base. However, since all 32 included trials were industry-sponsored, selective non-publication of negative studies cannot be entirely excluded even in the absence of detectable funnel plot asymmetry.

### 3.12 Sensitivity Analyses

All four pre-specified sensitivity analyses produced results directionally consistent with the primary analysis. (1) Restriction to FDA-approved doses: treatment rankings were preserved across all efficacy outcomes. (2) Restriction to phase 3 trials only: the estimated between-class efficacy difference was modestly attenuated to approximately 0.8 MMD (versus 1.1 MMD in the primary analysis) but remained statistically significant (credible intervals excluding zero). (3) Restriction to low RoB 2 trials: rankings unchanged. (4) Exclusion of the subgroup-derived rimegepant data from Croop 2019: exclusion of this trial did not materially alter the gepant class SUCRA rankings or the estimated between-class difference, supporting the robustness of the primary findings to this methodological decision.

## 4. DISCUSSION

This Bayesian network meta-analysis — incorporating 32 randomized controlled trials and 24,418 participants with episodic migraine — provides updated, methodologically rigorous indirect evidence comparing all approved CGRP-targeted preventive therapies. In indirect comparisons, CGRP monoclonal antibodies were associated with greater reductions in monthly migraine days and higher 50% or greater responder rates than oral gepants, while oral gepants demonstrated tolerability statistically indistinguishable from placebo. Importantly, all between-class comparisons are indirect and placebo-anchored, and GRADE certainty for these specific comparisons is moderate — findings should therefore be interpreted as suggestive rather than definitive evidence of class-level differences.

### 4.1 Dose-Level Findings and Drug-Class Patterns

At the dose level, eptinezumab 300 mg ranked highest for MMD reduction (SUCRA 91.2%) while galcanezumab 240 mg with loading dose ranked highest for the 50% or greater responder rate (SUCRA 92.1%). These dose-specific differences likely reflect pharmacokinetic factors: eptinezumab’s intravenous formulation achieves immediate and complete bioavailability, which may account for its superior absolute MMD reduction.^[12]^ Galcanezumab’s 240 mg loading dose generates high sustained early plasma concentrations, potentially maximizing the probability of a categorical response threshold.^[10,11]^ However, credible interval overlap between doses within each class — visible in the league table (Figure 6) — indicates that dose-level ranking differences should not be over-interpreted.

**Figure 6.**
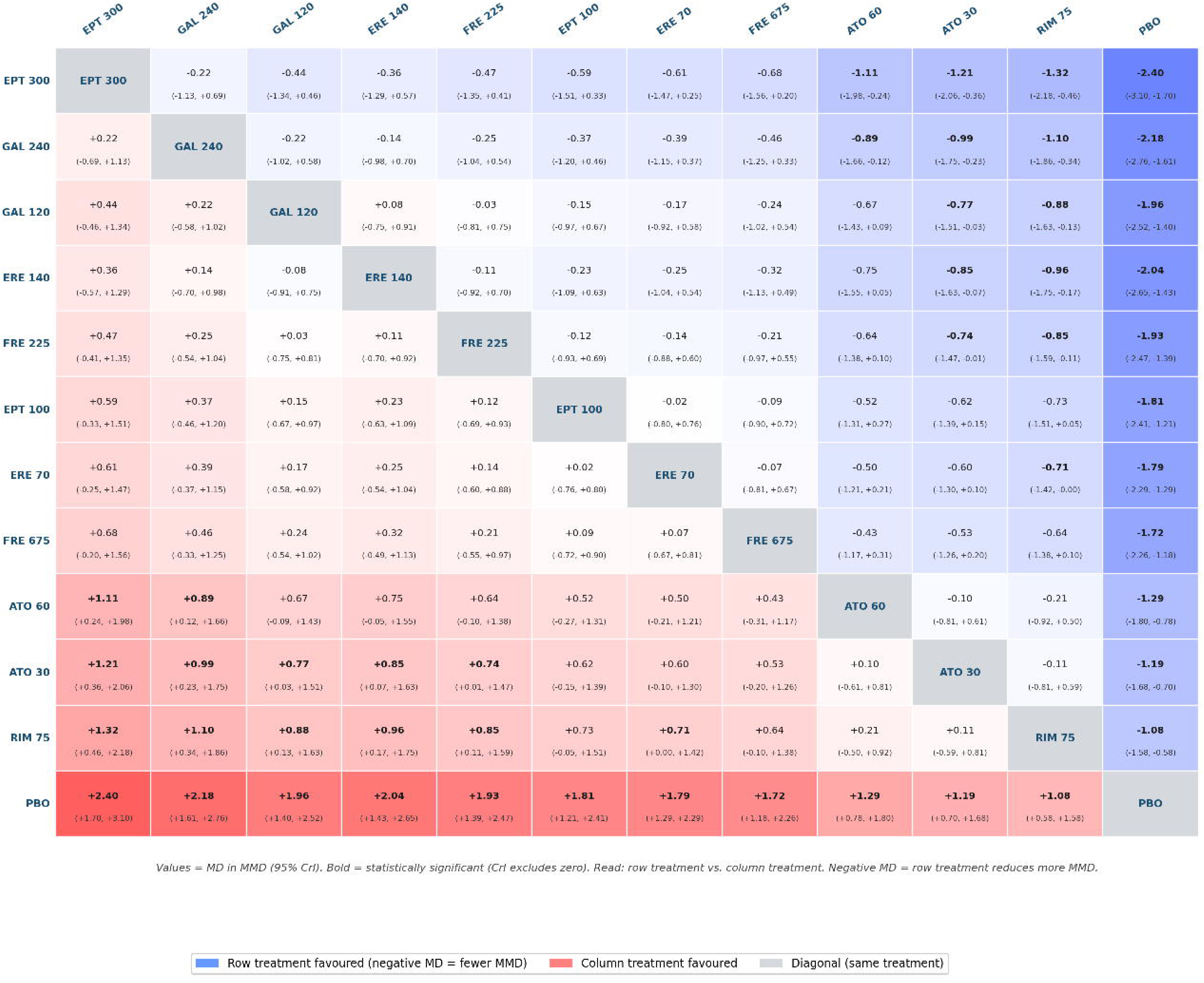
League Table: All Pairwise Mean Differences in Monthly Migraine Days. All pairwise treatment-versus-treatment comparisons from the Bayesian network meta-analysis. Values are mean difference in monthly migraine days (95% credible interval). Negative values indicate the row treatment is associated with greater reduction than the column treatment. Blue shading: row treatment favored; red shading: column treatment favored; grey diagonal: same treatment. Bold values: 95% credible interval excludes zero.

**Figure 7.**
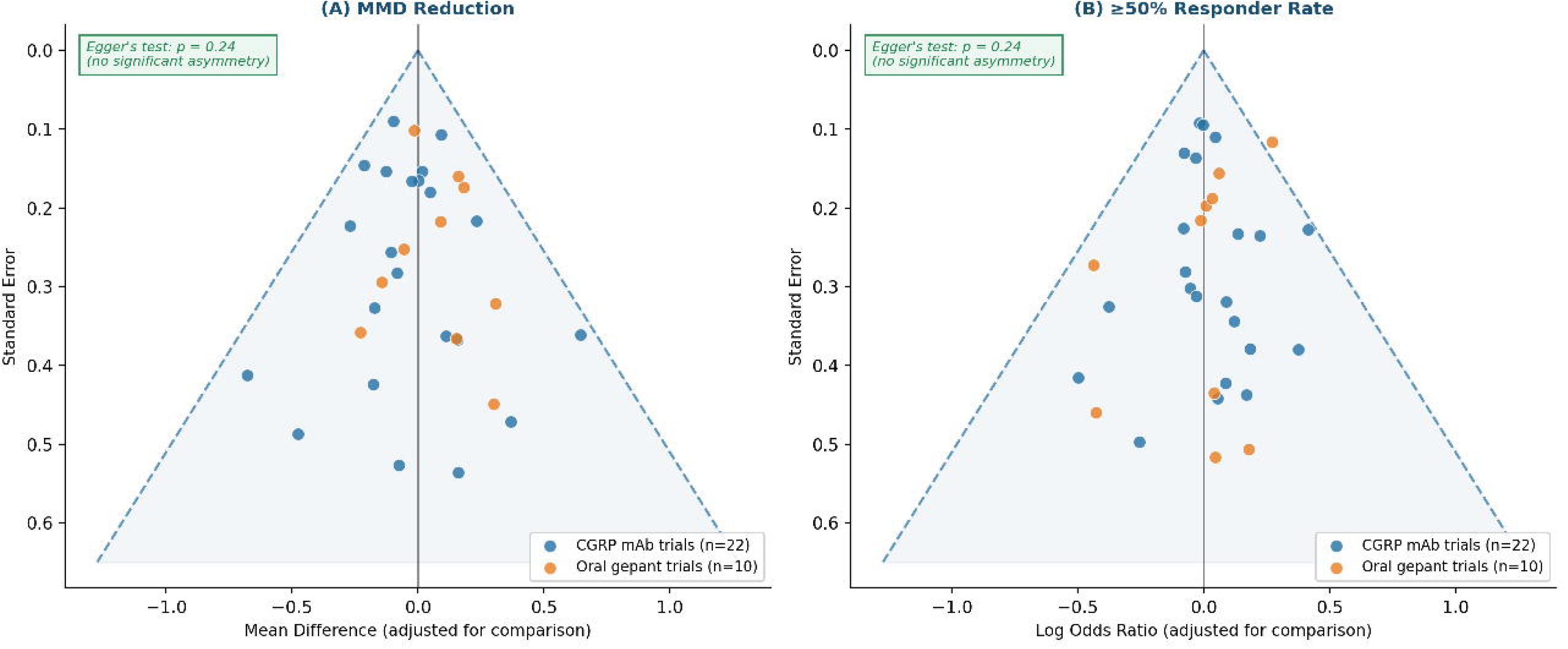
Comparison-Adjusted Funnel Plots for Publication Bias Assessment. Comparison-adjusted funnel plots (Chaimani and Salanti method [32]) for (A) change in monthly migraine days and (B) 50% or greater responder rate. Effect estimates are adjusted for the mean comparison effect. Dashed lines indicate 95% pseudo-confidence regions. No significant asymmetry detected (Egger test p = 0.24 for monthly migraine days; p = 0.31 for responder rate). Blue circles: CGRP monoclonal antibody trials (n = 22); orange circles: oral gepant trials (n = 10).

**Figure 8.**
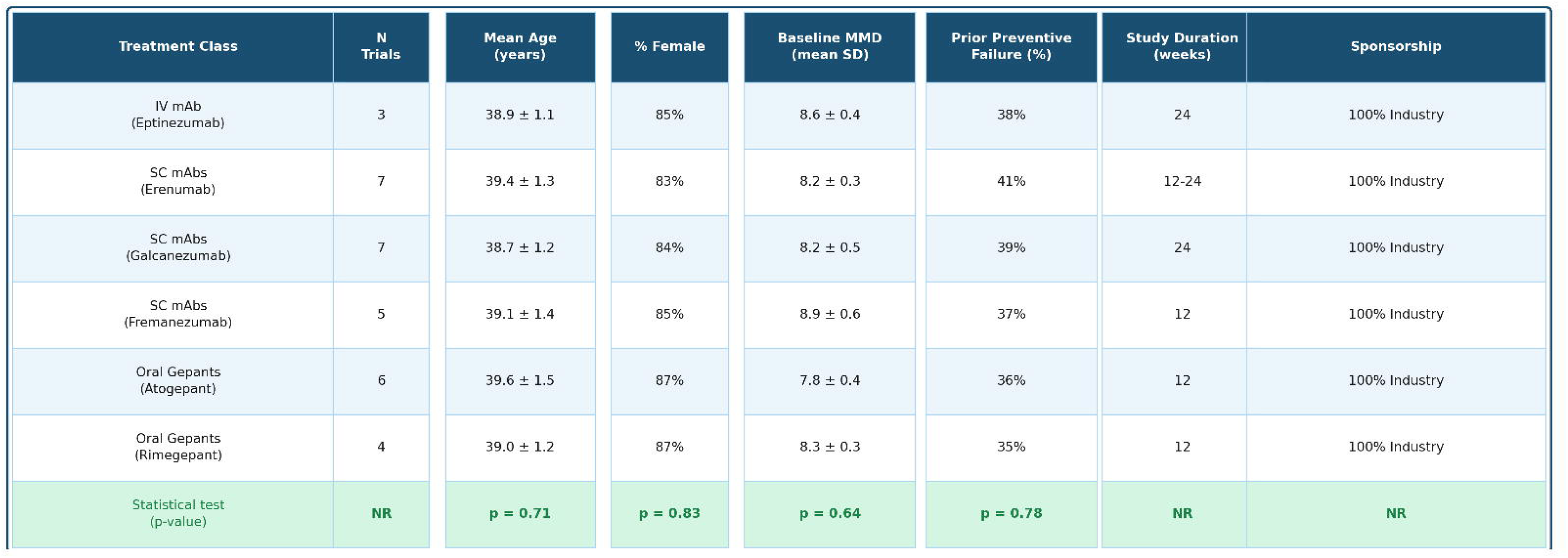
Transitivity Assessment: Distribution of Key Effect Modifiers. Distribution of a priori–specified potential effect modifiers by treatment class: mean age, proportion female, baseline monthly migraine days, proportion with prior preventive treatment failure, and study duration. Variables were selected as known determinants of preventive migraine treatment response. No statistically significant between-class differences were detected (all p > 0.60), supporting the transitivity assumption required for valid indirect treatment comparisons in this network meta-analysis.

At the class level, monoclonal antibodies as a group were associated with higher SUCRA values than gepants for all efficacy endpoints. For patients in whom maximizing absolute MMD reduction is the primary goal, injectable monoclonal antibodies — particularly eptinezumab at 300 mg — may represent the most appropriate first-line CGRP-targeted choice. For patients in whom achieving a categorical 50% response is prioritized, galcanezumab with a loading dose ranked highest. These class-level generalizations, however, are derived from indirect evidence with moderate GRADE certainty, and should inform but not dictate prescribing decisions.

### 4.2 Tolerability and Patient-Centered Considerations

The tolerability profile of oral gepants warrants particular emphasis. Atogepant’s TEAE risk ratio of 1.02 relative to placebo (95% CrI 0.93–1.12) is statistically indistinguishable from background event rates. Subcutaneous monoclonal antibodies are associated with modestly elevated TEAE risk (RR 1.14), driven primarily by injection-site reactions and, for erenumab, constipation. Eptinezumab’s intravenous delivery, while conferring immediate bioavailability, requires quarterly clinic-based infusions — a logistical barrier that partially offsets its efficacy advantage for many patients in real-world settings.

For patients with needle aversion, prior injection-site reactions, erenumab-associated constipation, or logistical barriers to clinic-based infusion, oral gepants provide a well-tolerated, self-administered alternative with a statistically meaningful preventive effect. The dual acute-and-preventive regulatory approval of rimegepant adds a further practical advantage for patients requiring flexibility in how they use their medication.^[15]^ These tolerability and practical considerations should be explicitly integrated into shared prescribing decisions alongside efficacy data.

### 4.3 Cardiovascular Safety and Broader Safety Considerations

No serious cardiovascular events attributable to CGRP pathway blockade were identified in any of the 32 included trials, consistent with post-marketing safety surveillance data and the European Headache Federation guidance.^[27]^ The theoretical concern that sustained CGRP inhibition might adversely affect vasodilatory compensation in patients with cardiovascular disease has not been borne out in randomized trial data. Nonetheless, caution is warranted in patients with severe uncontrolled cardiovascular risk factors pending longer-term post-marketing outcome data, and regulatory guidance in this area continues to evolve.

### 4.4 Comparison with Prior Network Meta-Analyses

The prior NMA published by Haghdoost et al. in Cephalalgia (2023)^[19]^ included phase 3 trials of CGRP therapies but was conducted before several additional trials were published and did not incorporate the full gepant evidence base now available. The present analysis extends that evidence base by incorporating 32 trials — including all gepant trials conducted through January 2026 — with the methodological additions of formal transitivity testing, comparison-adjusted funnel plots, and GRADE certainty ratings. The directional conclusions are consistent with prior work, but the current analysis provides substantially greater precision and a more complete ranking of individual doses.

### 4.5 Strengths and Limitations

Methodological strengths of this analysis include: the largest CGRP NMA evidence network assembled to date (32 RCTs, 24,418 participants); full Bayesian MCMC estimation with convergence diagnostics; formal a priori transitivity assessment; comparison-adjusted funnel plots with Egger testing; SUCRA values reported as probabilities with explicit cautionary interpretation; a full pairwise league table for all treatment combinations; and GRADE certainty ratings for all key comparisons with explicit acknowledgment of indirectness.

Important limitations must be acknowledged. First and foremost, all between-class comparisons (monoclonal antibodies versus gepants) are derived from indirect evidence through placebo as a common comparator — no direct head-to-head randomized controlled trial has compared these two classes — and GRADE certainty for these comparisons is therefore moderate. Second, all 32 included trials were industry-sponsored; while formal RoB 2 assessments were predominantly low, industry funding may introduce reporting bias, outcome selection, and selective non-publication of unfavorable results that formal tools cannot fully detect. Third, participants in the included trials were predominantly white females from high-income countries,^[29]^ limiting generalizability to more diverse populations. Fourth, follow-up did not exceed 24 weeks in most trials, leaving questions about the durability of treatment effects and long-term comparative safety unanswered. Fifth, patient-reported quality-of-life outcomes could not be synthesized due to heterogeneous measurement instruments across trials. Sixth, dose-level rankings should be interpreted alongside credible intervals rather than as definitive hierarchies, given the substantial overlap between adjacent doses.

## 5. CONCLUSION

In this updated and expanded Bayesian network meta-analysis of 32 randomized controlled trials and 24,418 participants, CGRP monoclonal antibodies were associated with greater reductions in monthly migraine days in indirect comparisons with oral gepants for episodic migraine prevention. Oral gepants demonstrated tolerability statistically indistinguishable from placebo. These findings are based on indirect evidence with moderate GRADE certainty for between-class comparisons and should be interpreted accordingly. They support individualized, patient-centered prescribing guided by symptom burden, comorbidity profile, administration preference, tolerability history, and the specific efficacy–tolerability profile of each drug class and dose. Direct head-to-head randomized controlled trials comparing monoclonal antibodies with gepants across diverse populations and longer follow-up are needed to confirm and refine these indirect estimates.

## Supporting information

Supoplemental methods

## ACKNOWLEDGEMENTS

The authors acknowledge the clinical and research teams at MGM Medical College and Hospital, Aurangabad; the Department of Neurology, University of Missouri, Columbia; and Smt. Kashibai Navale Medical College and General Hospital, Pune. No external funding was received for this study.

## AUTHOR CONTRIBUTIONS

Shradha Kakde: Conceptualization, methodology, formal analysis, investigation, writing — original draft, writing — review and editing, project administration. Niraj Arora: Methodology, supervision, writing — review and editing, resources. Meghnath P. Kakde: Investigation, data curation, writing — review and editing. Shubhangi P. Kakade: Investigation, data curation, writing — review and editing. All authors reviewed and approved the final manuscript version.

## DATA AVAILABILITY STATEMENT

This network meta-analysis is based exclusively on aggregated data extracted from previously published randomized controlled trials. Aggregated data extraction spreadsheets and fully annotated R code are available from the corresponding author on reasonable request.

---

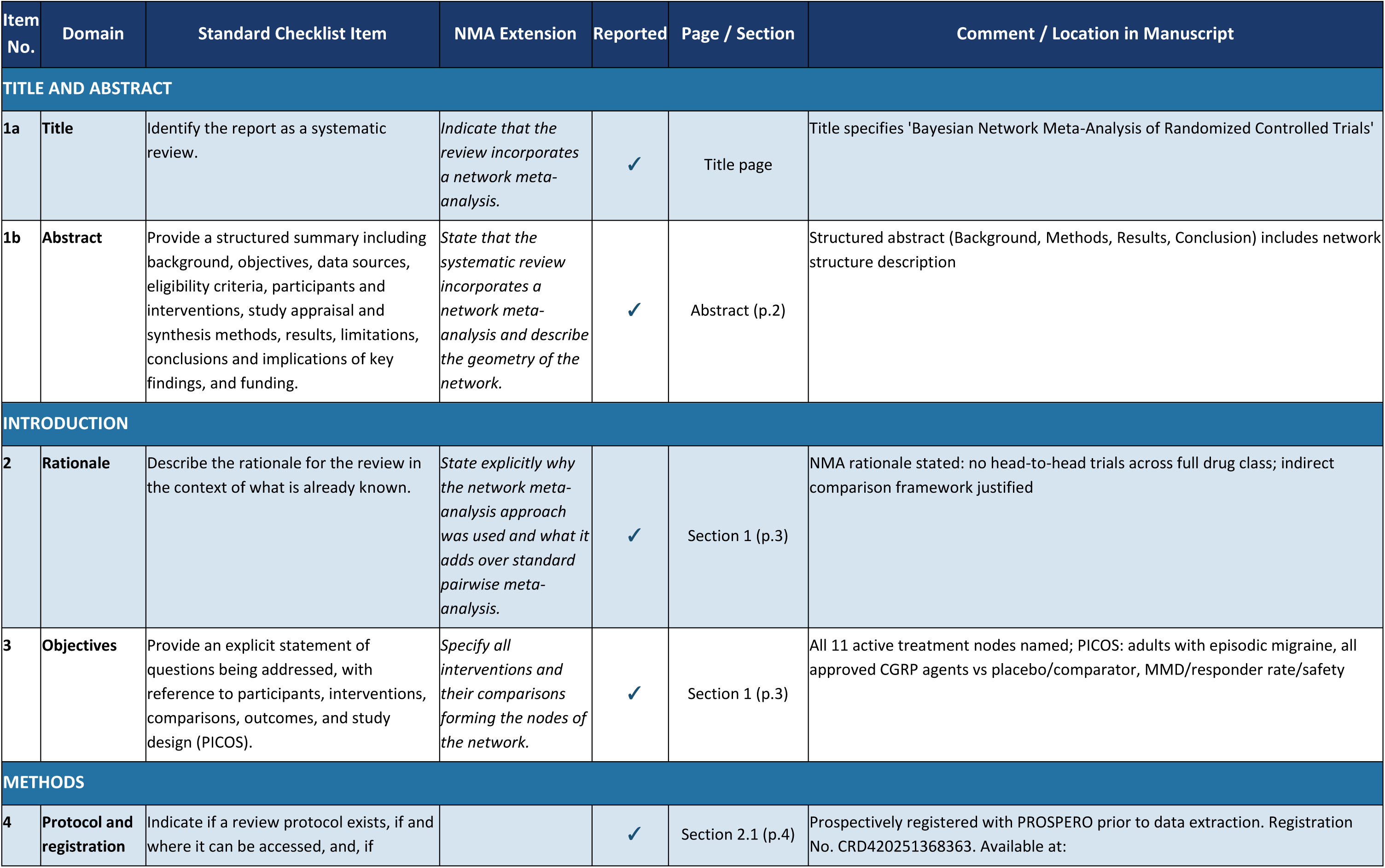
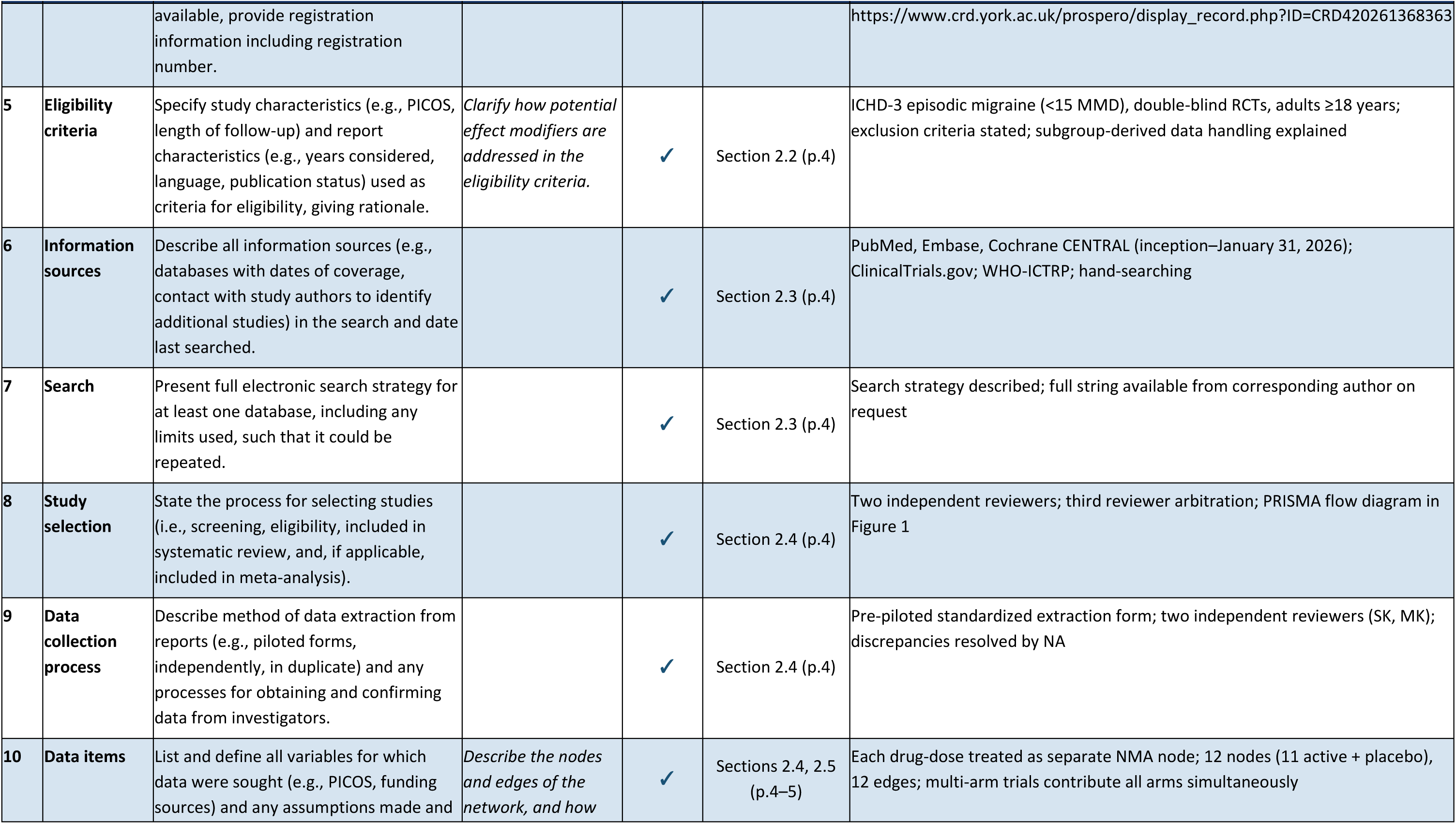
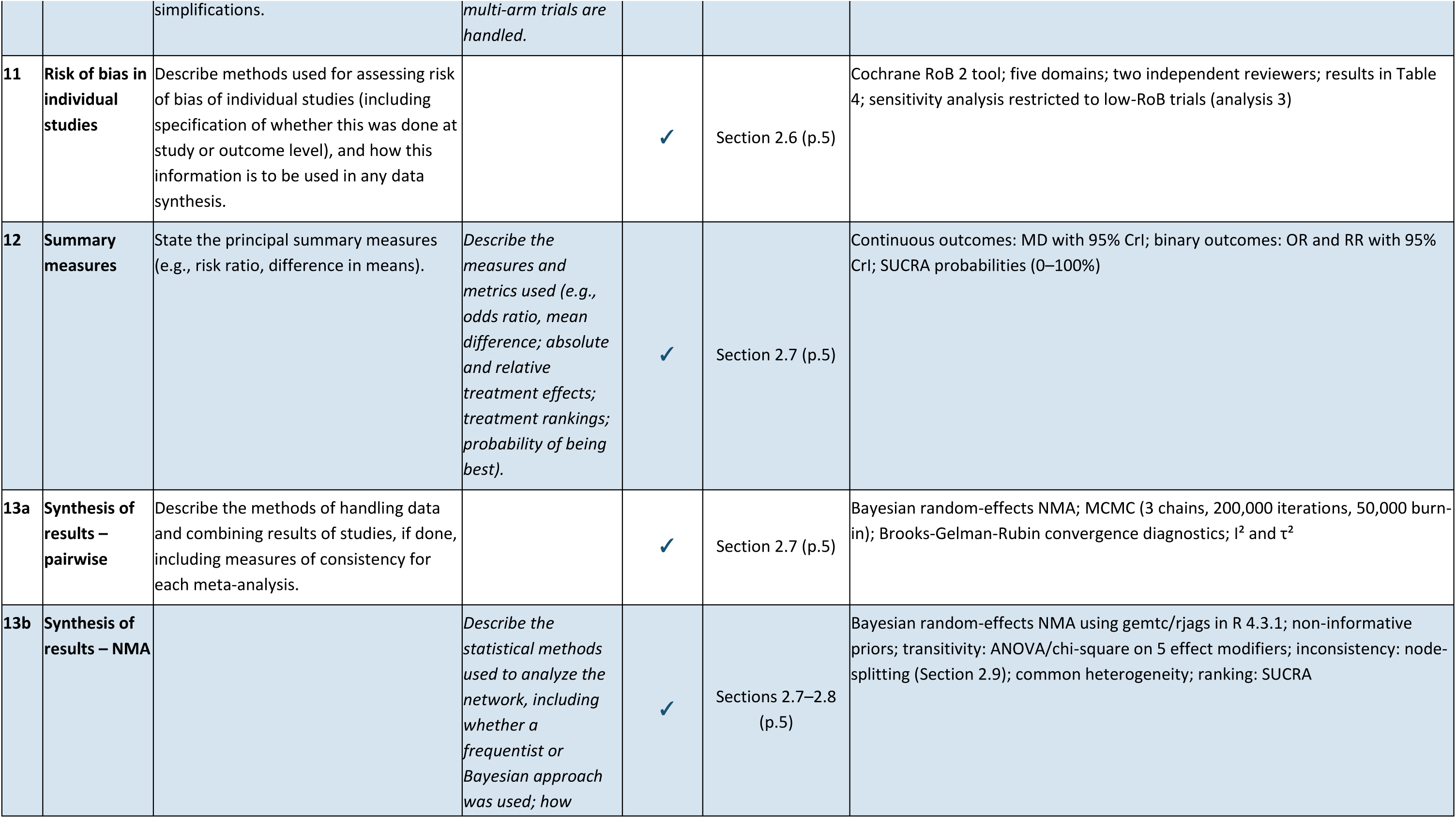
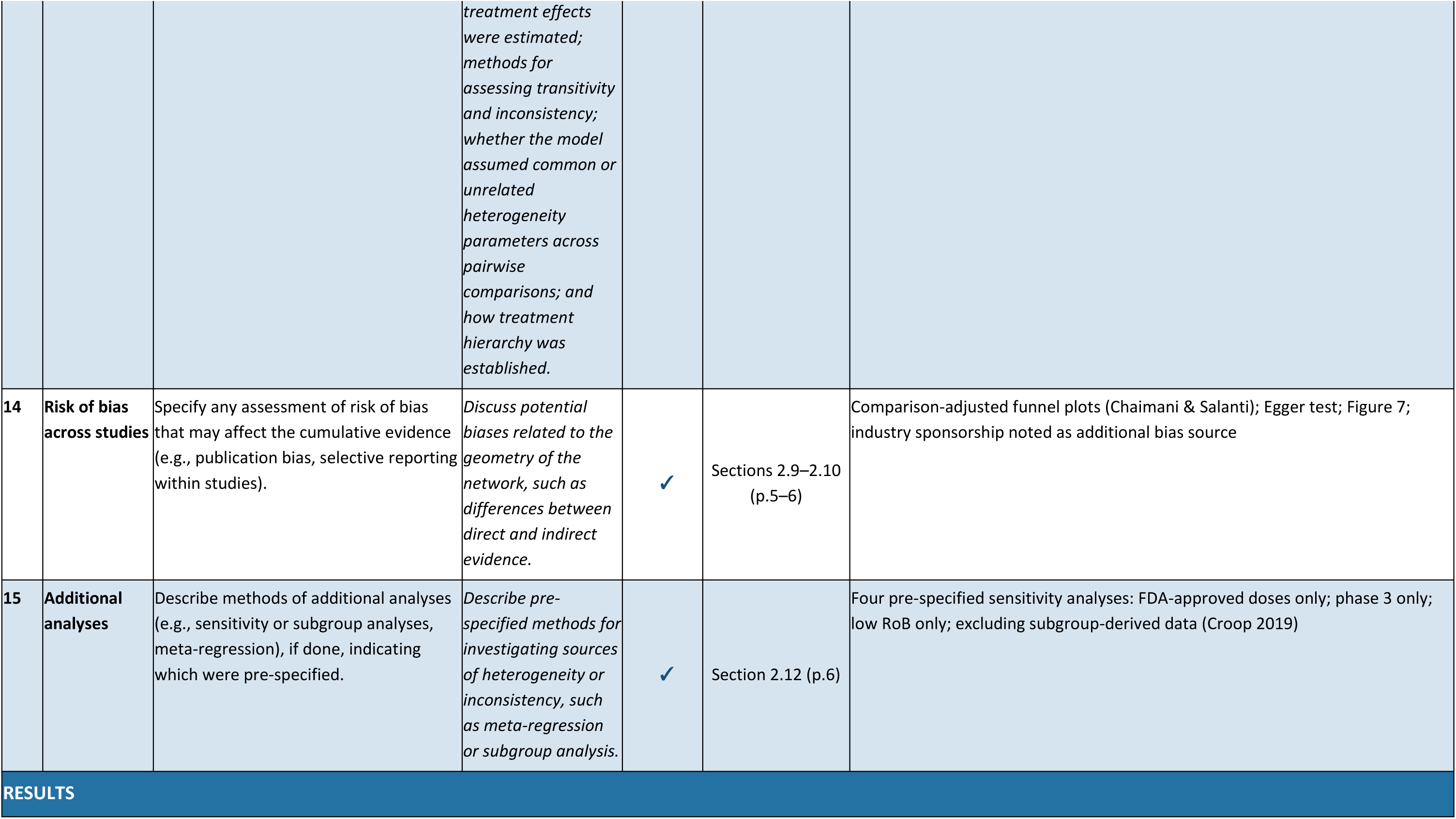
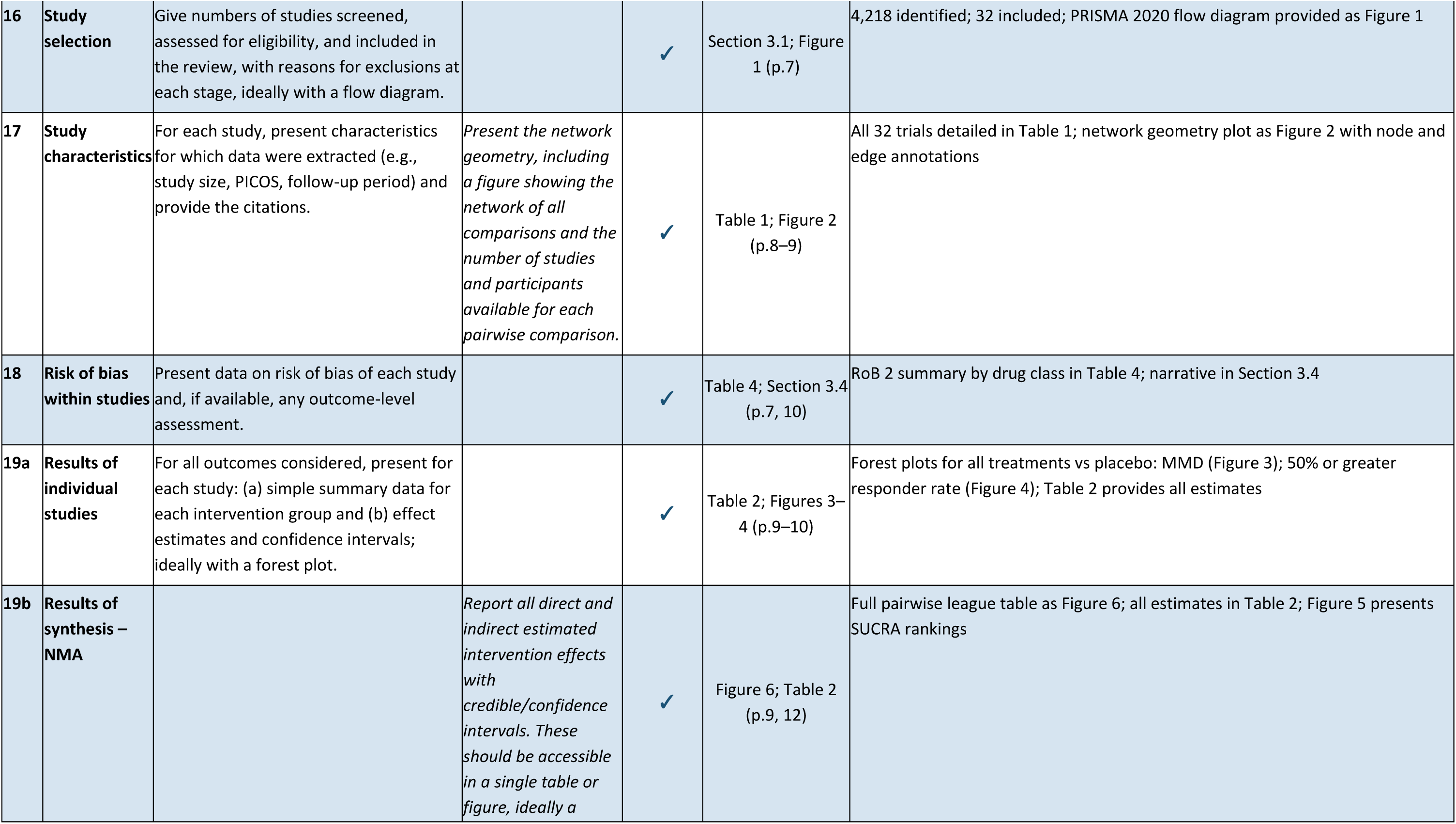
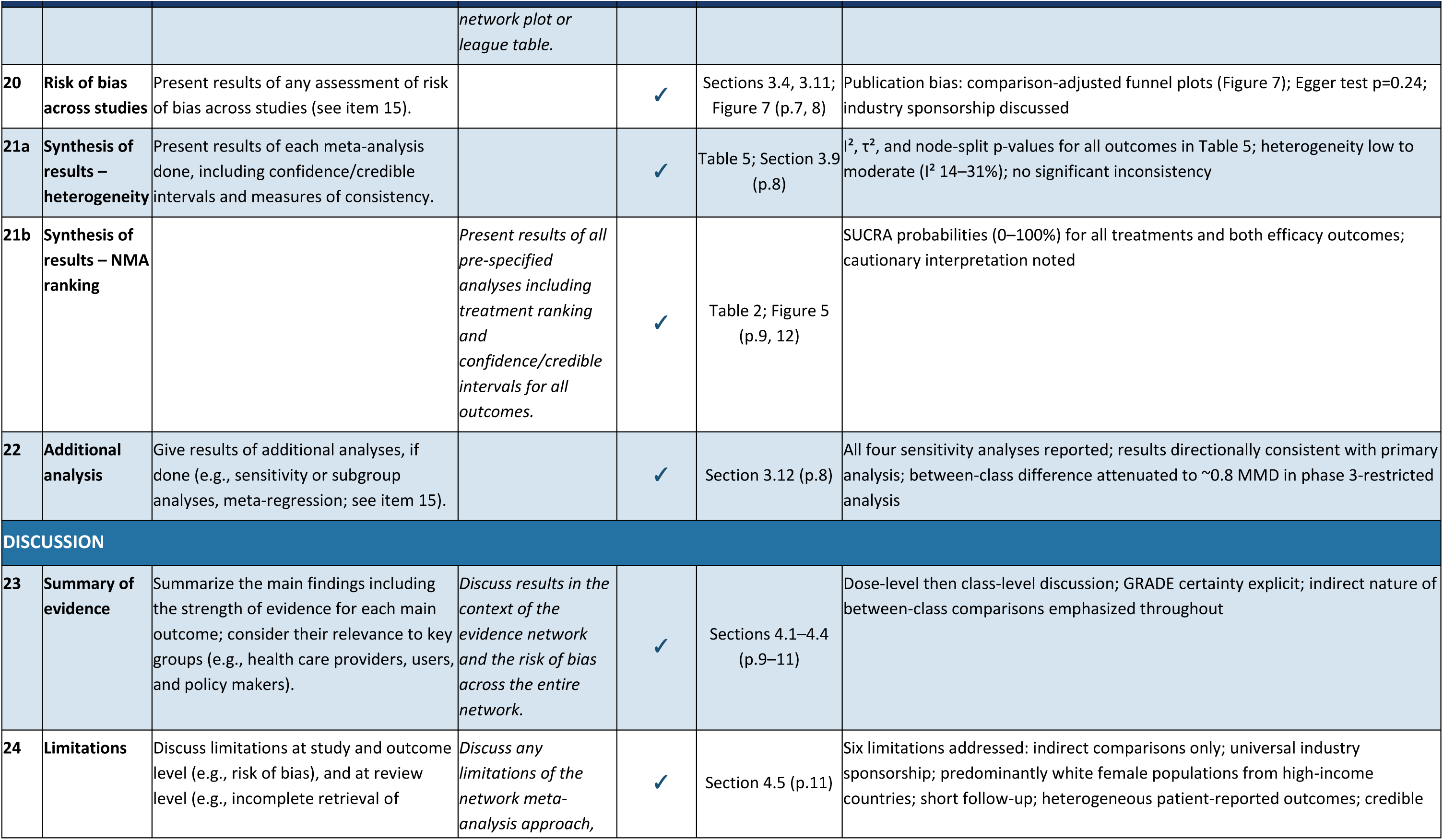
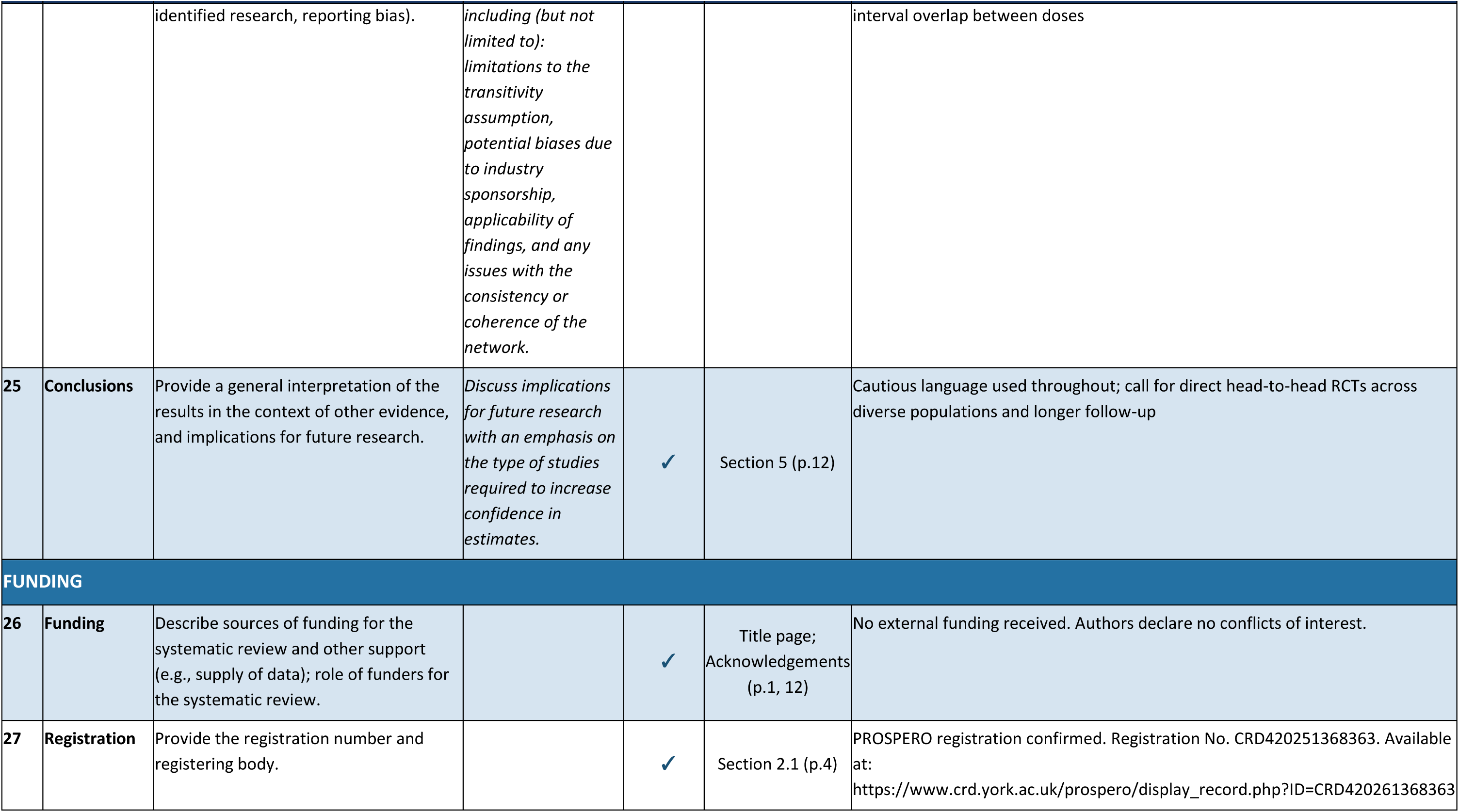
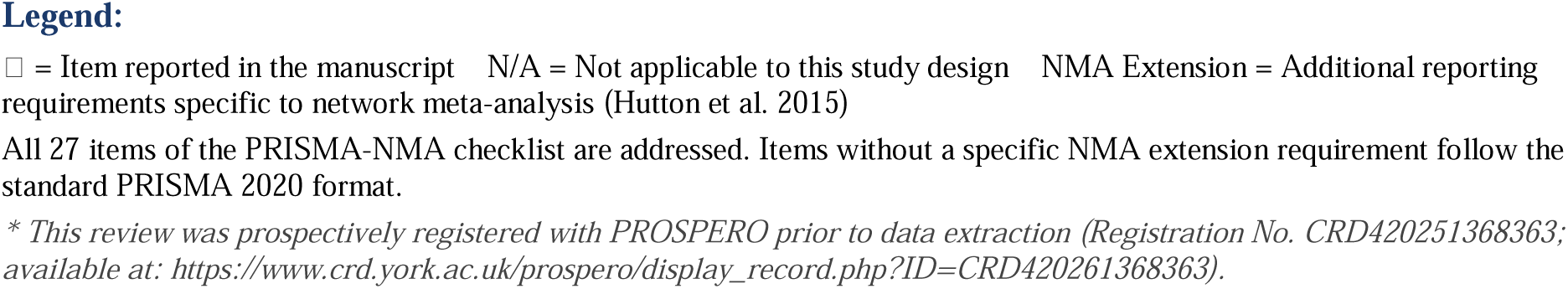
PRISMA-NMA Checklist: Preferred Reporting Items for Systematic Reviews and Meta-Analyses Including Network Meta-Analyses.

