## Supplementary material for "Comparative Efficacy and Safety of Calcitonin Gene-Related Peptide Monoclonal Antibodies Versus Oral Gepants for Episodic Migraine Prevention: A Bayesian Network Meta-Analysis of Randomized Controlled Trials": Supoplemental methods

### S1. Full Electronic Search Strategy

Databases searched: PubMed/MEDLINE, Embase, and Cochrane Central Register of Controlled Trials (CENTRAL). Search date: January 31, 2026. No language restrictions were applied. The following strategies were used:

**S1.1 PubMed/MEDLINE Search Strategy**

Database: PubMed (National Library of Medicine). Search date: January 31, 2026.

```
#1  "calcitonin gene-related peptide"[MeSH Terms]
#2  "calcitonin gene-related peptide"[Title/Abstract]
#3  CGRP[Title/Abstract]
#4  erenumab[Title/Abstract] OR AMG334[Title/Abstract] OR Aimovig[Title/Abstract]
#5  fremanezumab[Title/Abstract] OR TEV48125[Title/Abstract] OR Ajovy[Title/Abstract]
#6  galcanezumab[Title/Abstract] OR LY2951742[Title/Abstract] OR Emgality[Title/Abstract]
#7  eptinezumab[Title/Abstract] OR ALD403[Title/Abstract] OR Vyepti[Title/Abstract]
#8  atogepant[Title/Abstract] OR AGN241689[Title/Abstract] OR Qulipta[Title/Abstract]
#9  rimegepant[Title/Abstract] OR BHV3000[Title/Abstract] OR Nurtec[Title/Abstract]
#10 gepant[Title/Abstract] OR "CGRP antagonist"[Title/Abstract]
#11 "monoclonal antibody"[Title/Abstract] AND migraine[Title/Abstract]
#12 #1 OR #2 OR #3 OR #4 OR #5 OR #6 OR #7 OR #8 OR #9 OR #10 OR #11
#13 "migraine disorders"[MeSH Terms]
#14 migraine[Title/Abstract]
#15 "episodic migraine"[Title/Abstract]
#16 #13 OR #14 OR #15
#17 "randomized controlled trial"[Publication Type]
#18 "randomized controlled trials as topic"[MeSH Terms]
#19 randomized[Title/Abstract] OR randomised[Title/Abstract]
#20 "double-blind"[Title/Abstract] OR "placebo-controlled"[Title/Abstract]
#21 RCT[Title/Abstract]
#22 #17 OR #18 OR #19 OR #20 OR #21
#23 #12 AND #16 AND #22
```

Total retrieved: 1,847 records (prior to deduplication across databases).

**S1.2 Embase Search Strategy**

Database: Embase (Elsevier). Search date: January 31, 2026. Search via Embase.com interface.

```
#1  'calcitonin gene related peptide'/exp
#2  'calcitonin gene-related peptide':ab,ti
#3  'CGRP':ab,ti
#4  'erenumab':ab,ti OR 'AMG 334':ab,ti OR 'AMG334':ab,ti
#5  'fremanezumab':ab,ti OR 'TEV-48125':ab,ti OR 'TEV48125':ab,ti
#6  'galcanezumab':ab,ti OR 'LY2951742':ab,ti
#7  'eptinezumab':ab,ti OR 'ALD403':ab,ti
#8  'atogepant':ab,ti OR 'AGN-241689':ab,ti
#9  'rimegepant':ab,ti OR 'BHV-3000':ab,ti OR 'BHV3000':ab,ti
#10 'gepant':ab,ti OR 'CGRP receptor antagonist':ab,ti
#11 #1 OR #2 OR #3 OR #4 OR #5 OR #6 OR #7 OR #8 OR #9 OR #10
#12 'migraine'/exp
#13 'episodic migraine':ab,ti OR 'migraine prevention':ab,ti
#14 #12 OR #13
#15 'randomized controlled trial'/exp
#16 'randomization'/exp
#17 random*:ab,ti
#18 'double blind procedure'/exp
#19 'placebo'/exp AND 'controlled study'/exp
#20 #15 OR #16 OR #17 OR #18 OR #19
#21 #11 AND #14 AND #20
#22 #21 AND [humans]/lim
```

Total retrieved: 2,104 records (prior to deduplication across databases).

**S1.3 Cochrane CENTRAL Search Strategy**

Database: Cochrane Central Register of Controlled Trials (CENTRAL) via Cochrane Library. Search date: January 31, 2026.

```
#1  MeSH descriptor: [Calcitonin Gene-Related Peptide] explode all trees
#2  "calcitonin gene-related peptide" OR "CGRP":ti,ab,kw
#3  erenumab OR fremanezumab OR galcanezumab OR eptinezumab:ti,ab,kw
#4  atogepant OR rimegepant OR gepant:ti,ab,kw
#5  "monoclonal antibody" AND migraine:ti,ab,kw
#6  #1 OR #2 OR #3 OR #4 OR #5
#7  MeSH descriptor: [Migraine Disorders] explode all trees
#8  "episodic migraine" OR migraine:ti,ab,kw
#9  #7 OR #8
#10 #6 AND #9
```

Total retrieved: 267 records (CENTRAL, prior to deduplication). Cochrane CENTRAL already indexes RCTs exclusively; no additional RCT filter was applied.

**S1.4 ClinicalTrials.gov and WHO-ICTRP Search**

ClinicalTrials.gov (clinicaltrials.gov) was searched using the following terms in the condition/disease field: "migraine prevention" combined with interventions: "erenumab", "fremanezumab", "galcanezumab", "eptinezumab", "atogepant", "rimegepant", "CGRP". Filters applied: interventional studies; phase 2, 2/3, or 3; completed. Search date: January 31, 2026. Retrieved: 38 records.

The WHO International Clinical Trials Registry Platform (apps.who.int/trialsearch) was searched using the terms "migraine" AND "CGRP" AND "randomized". Retrieved: 9 records. These searches identified 47 additional records total beyond those obtained through database searching; no eligible unpublished trials meeting inclusion criteria were identified.

**S1.5 Deduplication**

Records retrieved: PubMed 1,847; Embase 2,104; Cochrane CENTRAL 267; ClinicalTrials.gov and WHO-ICTRP 47. Total: 4,265 records. After automated deduplication using Rayyan (for exact and fuzzy duplicates) and manual verification, 1,064 duplicate records were removed, yielding 3,201 unique records for title/abstract screening. The PRISMA 2020 flow diagram (Figure 1 of the main manuscript; reproduced as Figure S1 in this supplement) details all subsequent screening and selection steps.

### S2. Eligibility Criteria (PICOS Framework)

The following table summarizes all pre-specified inclusion and exclusion criteria organized by the PICOS framework. These criteria were finalized prior to data extraction and registered with PROSPERO.

| **PICOS Component** | **Inclusion Criteria** | **Exclusion Criteria** |
| --- | --- | --- |
| **Population (P)** | Adults ≥18 years with episodic migraine (<15 headache days/month) diagnosed per ICHD-3 criteria (Headache Classification Committee, 2018). No restriction on prior preventive treatment history, except as specified in sensitivity analyses. | Participants <18 years. Participants with chronic migraine only (≥15 headache days/month per ICHD-3) unless the study reported episodic migraine subgroup data separately, which were extracted per pre-specified protocol. Participants with a primary diagnosis of cluster headache, tension-type headache, or other non-migraine headache disorder (unless episodic migraine outcomes were separately reported). |
| **Intervention (I)** | Any regulatory-approved or clinically investigated dose of: (a) CGRP monoclonal antibody: erenumab (70 mg, 140 mg), fremanezumab (225 mg monthly, 675 mg quarterly), galcanezumab (120 mg, 240 mg with loading dose), eptinezumab (100 mg, 300 mg); or (b) Oral gepant: atogepant (10 mg, 30 mg, 60 mg), rimegepant (75 mg every other day). Each drug-dose combination treated as a separate NMA node. | Drugs not approved or investigated for preventive migraine use. Interventions outside the CGRP pathway (e.g., onabotulinumtoxinA, topiramate, valproate, beta-blockers). Acute-only dosing regimens (unless preventive outcome data were separately reported for a sub-cohort on regular preventive dosing). |
| **Comparator (C)** | Placebo (primary comparator, providing common network anchor for all indirect comparisons). Active comparator (other approved CGRP agents or doses, providing direct head-to-head evidence where available). Trials required at least a placebo arm. | Trials comparing only active agents versus active agents with no placebo arm and no connection to the placebo node in the network (would have disconnected the network graph and precluded indirect comparison). |
| **Outcomes (O)** | Primary: Mean change from baseline in monthly migraine days (MMD) at end of double-blind treatment period (weeks 12–24). Secondary: (1) 50% or greater responder rate (proportion achieving ≥50% reduction in MMD from baseline). (2) Incidence of treatment-emergent adverse events (TEAEs) — any-grade. (3) Rate of discontinuation due to adverse events (DAEs). All outcomes assessed at end of double-blind period. | Trials reporting outcomes only as absolute headache days without change-from-baseline data and where transformation was not possible. Trials reporting only patient-reported quality-of-life outcomes without any pre-specified MMD or responder rate endpoint (such trials could not contribute to the primary NMA; if they also reported MMD data, they were included). |
| **Study design (S)** | Double-blind, randomized controlled trials (phase 2b, 2/3, or phase 3). Parallel-group design. Minimum 12 weeks of double-blind treatment. Any publication status (including conference abstracts if sufficient data were available). Both published and unpublished registered trial data. | Open-label studies, open-label extension phases (unless baseline/double-blind data were separately extractable). Single-blind designs. Crossover trials without extractable parallel-group data for the first treatment period. Observational studies (cohort, case-control, cross-sectional). Case reports and case series. Reviews and meta-analyses. Sub-studies and post-hoc analyses of included trials (to avoid double-counting) unless they reported a distinct eligible subgroup not otherwise represented. Preclinical studies. |
| **Other eligibility criteria** | Trials must have contributed at least one extractable outcome (MMD, responder rate, TEAE, or DAE) at the pre-specified time point. English-language publications prioritized; non-English publications were included if sufficient data were reported in tables or figures for extraction. | Trials reporting exclusively on pediatric or adolescent populations (<18 years). Non-randomized studies of interventions (NRS). Unpublished registrations with no accessible trial data. Studies in migraine prophylaxis during pregnancy (population too distinct to contribute to the network). |

### S3. Data Extraction Sheet

The following standardized extraction form was piloted on five trials before use and was applied by two independent reviewers (SK and MK) to all 32 included trials. Discrepancies were resolved by a third reviewer (NA). This sheet represents the template used; actual extracted values for all 32 trials are available from the corresponding author on reasonable request.

| **Data Field** | **Details / Instructions** |
| --- | --- |
| **A. Study Identification** | |
| **Study ID / Reviewer initials** | Assign unique ID (e.g., STRIVE_2017). Record reviewer initials and extraction date. |
| **Full citation (Vancouver)** | Authors, title, journal, year, volume, pages, DOI. |
| **ClinicalTrials.gov / other registry number** | NCT number or equivalent registry identifier. |
| **Trial name / acronym** | e.g., STRIVE, ARISE, EVOLVE-1, ADVANCE, PROMISE-1. |
| **Funding source** | Industry-funded; government-funded; academic/no external funding; mixed. Note specific funder(s). |
| **Publication status** | Peer-reviewed full text; conference abstract only; other. |
| **B. Study Design** | |
| **Study design** | Double-blind RCT (parallel); crossover (if extractable); other. |
| **Phase** | Phase 2; Phase 2b; Phase 2/3; Phase 3; Phase 3b. |
| **Total double-blind treatment duration (weeks)** | Report in weeks (e.g., 12; 24). Note if different time points reported. |
| **Primary efficacy endpoint time point** | Week at which primary outcome is assessed (e.g., Week 12, Weeks 13–24). |
| **Geographic location of study** | Country/countries; multinational. |
| **C. Participant Characteristics** | |
| **Total randomized (N)** | Total randomized across all arms. |
| **N per arm (active / placebo)** | Provide separately for each arm. |
| **Mean age (years) ± SD** | Pooled or per arm; note if median reported instead. |
| **Sex: % female** | Pooled across arms; per arm if substantially different. |
| **Migraine type** | Episodic (<15 MMD/month); chronic; mixed (if episodic subgroup extracted, note this). |
| **Mean baseline MMD ± SD** | As reported; note if different across arms. |
| **Proportion with prior preventive treatment failure (≥1 agent)** | %; note definition used (e.g., ≥1 prior failed preventive; ≥2). |
| **Proportion using acute medication overuse at baseline** | %; if reported. |
| **Duration of migraine history (years)** | Mean ± SD if reported. |
| **ICHD diagnostic criteria version used** | ICHD-2; ICHD-3; ICHD-3 beta; other. |
| **D. Intervention Details** | |
| **Drug name** | Generic name (e.g., erenumab, atogepant). |
| **Drug class** | CGRP monoclonal antibody (anti-CGRP ligand; anti-CGRP receptor); oral gepant (CGRP receptor antagonist). |
| **Dose(s) and frequency** | e.g., 70 mg subcutaneous monthly; 60 mg oral daily. List all active arms. |
| **Route of administration** | Subcutaneous (SC); intravenous (IV); oral. |
| **Loading dose used?** | Yes/No; if yes, specify. |
| **Comparator (control arm)** | Placebo; active comparator (specify); both. |
| **NMA node assignment** | Assign each arm to its pre-specified NMA node label (e.g., "Erenumab 140 mg"). |
| **Multi-arm trial?** | Yes/No; if yes, list all arms to be included simultaneously in NMA. |
| **E. Primary Outcome: Monthly Migraine Days (MMD)** | |
| **Mean change from baseline in MMD (active arm)** | Provide mean ± SD (or SE) at the pre-specified time point. Note time point. |
| **Mean change from baseline in MMD (placebo arm)** | Provide mean ± SD (or SE). Note time point. |
| **Between-group MD (active vs placebo) and 95% CI** | If directly reported; otherwise compute from arm-level data. |
| **P-value for primary outcome** | As reported. |
| **F. Secondary Outcome: 50% or Greater Responder Rate** | |
| **% achieving ≥50% MMD reduction (active arm)** | At pre-specified time point; note time point. |
| **% achieving ≥50% MMD reduction (placebo arm)** | At pre-specified time point. |
| **OR or RR with 95% CI (if reported)** | As reported; otherwise extracted for computation. |
| **P-value** | As reported. |
| **G. Safety Outcomes** | |
| **Any TEAE — % active arm** | Total treatment-emergent adverse event rate, any grade, at any point during double-blind period. |
| **Any TEAE — % placebo arm** | As above for placebo. |
| **Discontinuation due to AE — % active arm** | Premature discontinuation attributed to adverse event. |
| **Discontinuation due to AE — % placebo arm** | As above for placebo. |
| **Serious adverse events — % active arm** | If reported. |
| **Serious adverse events — % placebo arm** | If reported. |
| **Notable drug-specific adverse events** | e.g., injection-site reaction, constipation (erenumab), nasopharyngitis (eptinezumab), nausea (atogepant). Provide % if reported. |
| **Cardiovascular adverse events** | Rate of serious CV events; type; attribution to CGRP pathway. |
| **H. Risk of Bias Assessment (Cochrane RoB 2)** | |
| **Domain 1: Randomization process** | Low / Some concern / High. Rationale. |
| **Domain 2: Deviations from intended interventions** | Low / Some concern / High. Rationale. |
| **Domain 3: Missing outcome data** | Low / Some concern / High. Rationale. |
| **Domain 4: Measurement of the outcome** | Low / Some concern / High. Rationale. |
| **Domain 5: Selection of the reported result** | Low / Some concern / High. Rationale. |
| **Overall RoB 2 rating** | Low / Some concern / High. |
| **Reviewer notes** | Any additional methodological concerns not captured by RoB 2 domains. |
| **I. Additional / Transitivity Variables** | |
| **Mean study duration (weeks)** | Double-blind treatment period. |
| **Mean age (pooled across arms, years)** | For transitivity analysis. |
| **Proportion female (pooled, %)** | For transitivity analysis. |
| **Mean baseline MMD (pooled)** | For transitivity analysis. |
| **Proportion with prior preventive failure (%)** | For transitivity analysis. |
| **Data source for extractable subgroup** | Applicable to trials contributing subgroup data only (e.g., Croop 2019 rimegepant preventive analysis): note specific table/figure from source publication. |

### S4. PRISMA 2020 Flow Diagram (Figure S1)

The following table presents the PRISMA 2020 flow diagram in tabular format, showing the number of records at each stage of study selection. This corresponds to Figure 1 of the main manuscript.

| **Stage** | **n** | **Notes** |
| --- | --- | --- |
| **IDENTIFICATION** | | |
| Records identified via PubMed (inception–January 31, 2026) | **1,847** |  |
| Records identified via Embase (inception–January 31, 2026) | **2,104** |  |
| Records identified via Cochrane CENTRAL (inception–January 31, 2026) | **267** |  |
| Records retrieved from ClinicalTrials.gov | **38** |  |
| Records retrieved from WHO-ICTRP | **9** |  |
| TOTAL records identified | **4,265** |  |
| Duplicate records removed (Rayyan automated + manual verification) | **1,064** |  |
| Records remaining after deduplication | **3,201** |  |
| **SCREENING** | | |
| Records screened (title and abstract) | **3,201** |  |
| Records excluded at title/abstract screening | **2,889** | Wrong study design (n=1,044); wrong population (n=712); not CGRP-targeted therapy (n=688); wrong outcome (n=445) |
| Full-text articles assessed for eligibility | **312** |  |
| Full-text articles excluded, with reasons: | **280** |  |
| — Wrong study design (open-label, non-RCT) |  | n = 68 |
| — Exclusively chronic migraine population, no episodic subgroup |  | n = 54 |
| — Duplicate dataset (same trial, different publication) |  | n = 42 |
| — Wrong intervention / dose not meeting inclusion threshold |  | n = 38 |
| — Insufficient data for extraction (abstract only, no extractable outcome) |  | n = 31 |
| — Open-label extension phase only |  | n = 24 |
| — Wrong comparator (no placebo arm, network disconnect) |  | n = 14 |
| — Follow-up <12 weeks |  | n = 9 |
| **INCLUSION** | | |
| Studies included in qualitative synthesis (systematic review) | **32** |  |
| — CGRP monoclonal antibody trials |  | n = 22 |
| — Oral gepant trials |  | n = 10 |
| Studies included in quantitative synthesis (Bayesian NMA) | **32** | All 32 contributed at least one extractable NMA outcome |
| Total participants randomized across included trials | **24,418** | Range per trial: 246–1,351 |

Abbreviations: CENTRAL, Cochrane Central Register of Controlled Trials; NMA, network meta-analysis; RCT, randomized controlled trial; WHO-ICTRP, World Health Organization International Clinical Trials Registry Platform.

Note: The PRISMA 2020 Statement (Page MJ et al. BMJ. 2021;372:n71) and the PRISMA-NMA extension (Hutton B et al. Ann Intern Med. 2015;162:777–784) were followed in preparing this flow diagram.
